# Dynamic Clinical Success Rates for Drugs in the 21st Century

**DOI:** 10.1101/2024.02.26.24303388

**Authors:** Ying Zhou, Yintao Zhang, Zhen Chen, Shijie Huang, Yinghong Li, Jianbo Fu, Hongning Zhang, Donghai Zhao, Xichen Lian, Yuan Zhou, Xinyi Shen, Yunqing Qiu, Lianyi Han, Feng Zhu

**Affiliations:** College of Pharmaceutical Sciences, Zhejiang University, Hangzhou 310058, China; Centre for Clinical Research, Helmholtz Centre for Infection Research, Hannover 30625, Germany; Yale School of Public Health, Yale University, New Haven 06510, United States; Greater Bay Area Institute of Precision Medicine, Fudan University, Shanghai 315211, China

## Abstract

In clinical drug development, two fundamental questions remain unanswered: what is the success rate of drugs in clinical trial? how does such rate change over time? Herein, a systematic analysis on the dynamic change of drugs’ *clinical success rates* (ClinSRs) using data from 20,398 clinical trial pipelines of 9,682 unique molecular entities during the past two decades was presented. Our analysis discovered that ClinSRs had been declining since the beginning of 21st century, and hit the bottom in recent years even substantially lower than previous estimates. In-depth assessments further reported great variation among the ClinSRs of various diseases, developmental strategies, and drug modalities. A platform ‘*ClinSR.org*’ (http://ClinSR.idrblab.org/) was finally constructed online to enable the illustration of how ClinSR dynamically changes over time, automated update of ClinSR for the coming decade, and customized calculation of ClinSRs for any drug group of interest. In sum, this study met the critical demand for accurate, timely and persistent assessment of ClinSR, for now and the future, to aid pharmaceutical and economic decision making.

## 1. Introduction

Drug discovery is characterized by a high attrition rate with limited approvals each year (1). For clinical drug development, it is critical to answer two fundamental questions: *What is the success rate of drugs in clinical trial*? (2), and *How do such rates change over time*? (3). The answers to these questions are crucial for both clinical researcher and pharmaceutical investor when making scientific and economic decision (4). As described in **Supplementary Table S1**, researchers have been working on addressing these questions during the past two decades. Some assess the clinical success rate of the entire pharmaceutical industry within a specific time-window (2–5), and others concentrate on particular therapeutic area or focus on individual disease indication (6–8).

However, there is a huge variation, ranging from 7% to 20%, in the reported *clinical success rate* (ClinSR) among previous studies (2–5), which may result from the heterogeneity of accumulated data, difference in evaluating protocols, and shift of studied time-windows. In other words, direct comparison among previously-reported ClinSRs can provide limited insight into how investment and technology affect the success of drug discovery (2–5), and a unified standard for clinical data collection and success rate evaluation is thus highly demanded. <u>Moreover</u>, due to the *lag of time* and *termination* in data collection, it is difficult for previous studies to timely report the ClinSRs of their publication year, and it is impossible for them to update the ClinSR for the coming decade (2–8). Therefore, it is of great interest to develop new strategy enabling timely and persistent data collection and automated assessment of the latest ClinSRs for any studied group of drugs.

In this study, a systematic analysis on dynamic *clinical success rate* (ClinSR) of drugs in the 21st century was therefore conducted. *First*, a strict and reproducible process for data collection and ClinSR evaluation was constructed, which worked out the dynamic shift (from 2001 to 2023) of ClinSRs using 20,398 clinical trial pipelines of 9,682 unique molecular entities. *Second*, in-depth evaluations of ClinSR were performed from diverse perspectives (such as various disease classes, distinct developmental strategies and different drug modalities), which provided concrete insight into particular directions of current pharmaceutical research. *Finally*, a multi-functional platform ClinSR.org was developed online (http://ClinSR.idrblab.org/) to enable the dynamic illustration of how ClinSR change over time, realize the automated update of ClinSRs for the coming decade, and allow the customized evaluation of ClinSR for any drug group of interest. In conclusion, this study could persistently support the pharmaceutical decision making for now and the future.

## 2. Material and Methods

### 2.1 Collection of Clinical Trial Drugs and Their Time-dependent Clinical Status

Data collection in this study consisted of two sequential procedures: (*a*) the accumulation of drug data from exiting databases, and (*b*) the data standardization facilitating subsequent analysis.

#### 2.1.1 Collection of Drug Data from Established Databases

Comparing with other established databases, *ClinicalTrials.gov* had long been considered as one of the most influential sources of clinical trial drugs and clinical testing information, which had rapidly expanded since 2007 due to the official supports from U.S. FDA (*2007 FDA Amendments Act* required all clinical trials to be registered into *Clinicaltrials.gov*). In this study, to ensure the reliability of clinical information and maintain the high criteria of data inclusion across different years, *ClinicalTrials.gov* was adopted as the only resource for collecting the data of clinical trial drugs. Moreover, the data of approved drugs were directly collected from the official website of U.S. FDA (https://www.fda.gov/drugs), which resulted in a total of 824 unique molecular entities approved between 2000 and 2023. As shown in **Figure 1**, the numbers of *New Drug Applications* (NDA colored in blue) and *Biologics License Applications* (BLA colored in pink) approved each year were provided. A unique molecular entity could be further approved for a new indication (a successful drug-repurposing) after its first approval. For example, *alemtuzumab* (as described in **Figure 1**) was first approved in 2001 for treating *B-cell chronic lymphocytic leukemia*, and later approved in 2014 for *multiple sclerosis*. Although *alemtuzumab* was not considered in this study as a newly approved BLA for 2014, it had been included as an approved drug-disease project in 2014. Therefore, the total number of approved drug-disease projects was illustrated in **Figure 1** (grey dash line), which was higher than the summation of NDAs and BLAs newly approved each year. These results indicated that drug-repurposing was quite active during the past two decades. Another two reputable databases included here for collecting drug information were *Therapeutic Target Database* (9) and *DrugBank* (10), which facilitated this study to further confirm the drug modality (such as: small molecular drug and antibody), key pharmaceutical and physicochemical characteristics (such as: molecular weight, logP, and structure), and so on. Such data were critical for ensuring customized analysis of ClinSR for particular groups of clinical trial drugs.

**Figure 1.**
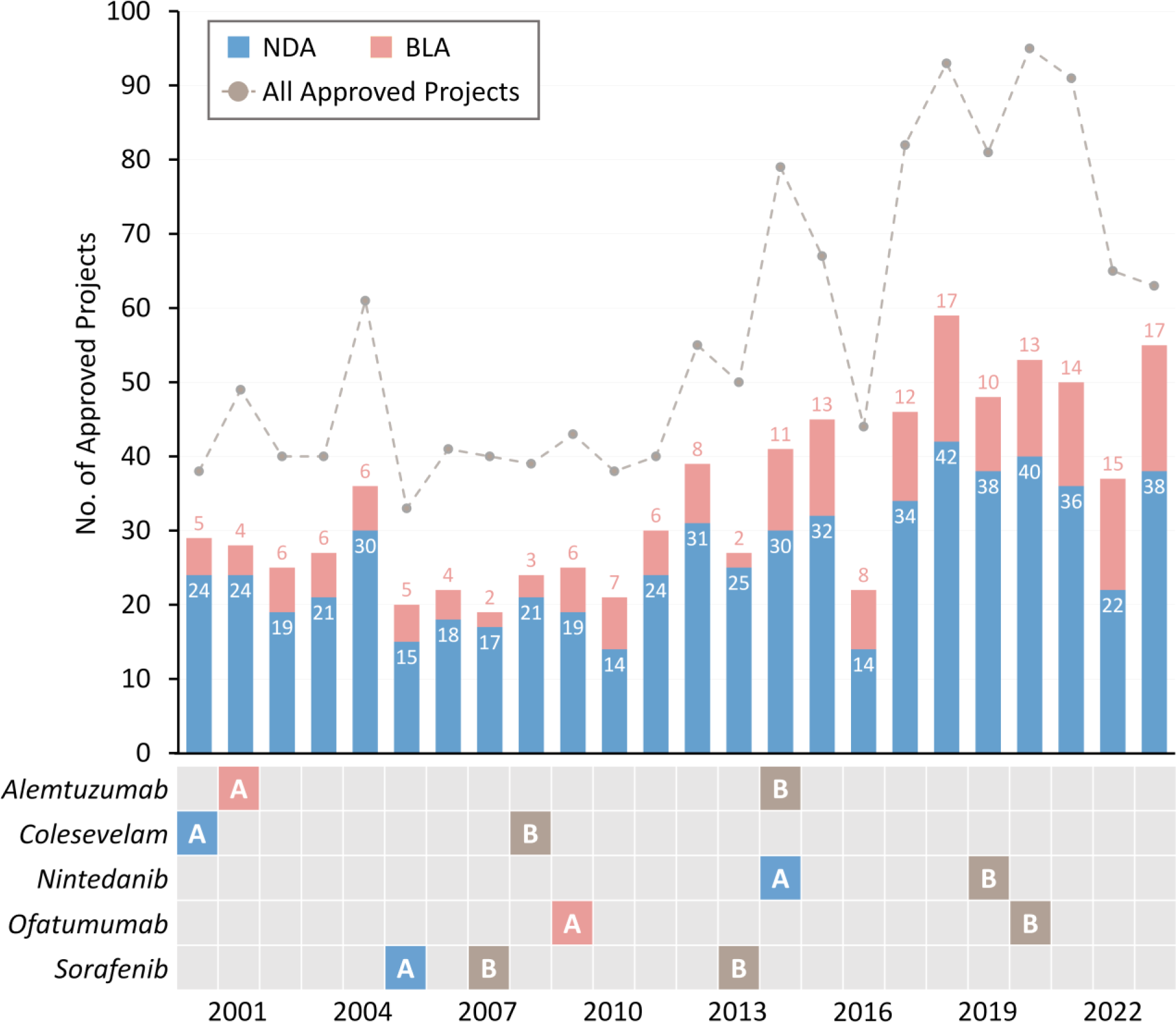
The numbers of *New Drug Applications* (NDA, colored in blue) and *Biologics License Applications* (BLA, colored in pink) approved by the US FDA each year between 2000 and 2023, inclusive. A unique molecular entity could be further approved for a new indication (a successful drug-repurposing) after its first approval. For example, *alemtuzumab* was first approved in 2001 for the treatment of *B-cell chronic lymphocytic leukemia*, and later in 2014 for *multiple sclerosis*. Although *alemtuzumab* was not considered in this study as a newly approved BLA for 2014, it had been included as an approved drug-disease project in 2014. Therefore, a grey dash line was provided to illustrate the total number of approved drug-disease projects, which was larger than the summation of NDAs and BLAs approved each year. ‘**A**’ indicated the first approval of a new molecular entity (blue for NDA, red for BLA), while ‘**B**’ indicated the approval of a repurposing indication after the initial approval of the corresponding unique molecular entity.

#### 2.1.2 Data Standardization for the Drugs in Clinical Trial

Clinical trial drug data were collected from the latest version (Jan 1st, 2024) of *ClinicalTrials.gov* as described above. To make it usable for success rate analysis, several data standardization steps were sequentially applied. *First*, many trials were excluded from this study, such as the ones with no clinical status described, the ones without clear time of trials, and the ones with no drug tested (such as: dental implant, liver transplant, and aerobic exercise). *Second*, detailed information for each clinical trial drug was systematically collected, which included clinical trial ID, drug name, developmental status (such as: Phase 1 and Phase 2/3), disease indication, date of trial start, date of study completion, recruitment status, and so on. *Third*, duplicate drug names were merged by adopting various existing synonym databases (such as PubChem, ADCdb, DrugBank, and TTD), and different formulations for an active pharmaceutical ingredient were also merged in this study. *Finally*, based on the method used in the pioneer study (2), the trials in Phase 1/2 were considered as Phase 2, and the trials in Phase 2/3 were regarded to be in Phase 3 in this study, and all diseases were standardized using the latest WHO *International Classification of Diseases* (11).

#### 2.1.3 Pipeline Identification for a Drug of Distinct Disease

The clinical trial pipeline (CTP) of a drug for treating one disease was generated in this study by merging all trials of this drug treating the same disease, and those trials of this drug treating other diseases were used to generated new CTPs. As described in **Figure 2*a*** (taking the drug *vilaprisan* as an example), it had been tested in clinical trials for two disease indications (*endometriosis* and *uterine leiomyoma*), which resulted in two distinct CTPs for this specific drug. As a result, a total of 20,398 CTPs corresponding to 9,682 unique molecular entities for treating 910 disease classes defined by the WHO ICD-11 (*acute myeloid leukemia*, *cholera*, *hyperlipoproteinemia*, *migraine*, *thalassaemias*, *etc.*) were collected for the subsequent analysis of clinical success rate.

**Figure 2.**
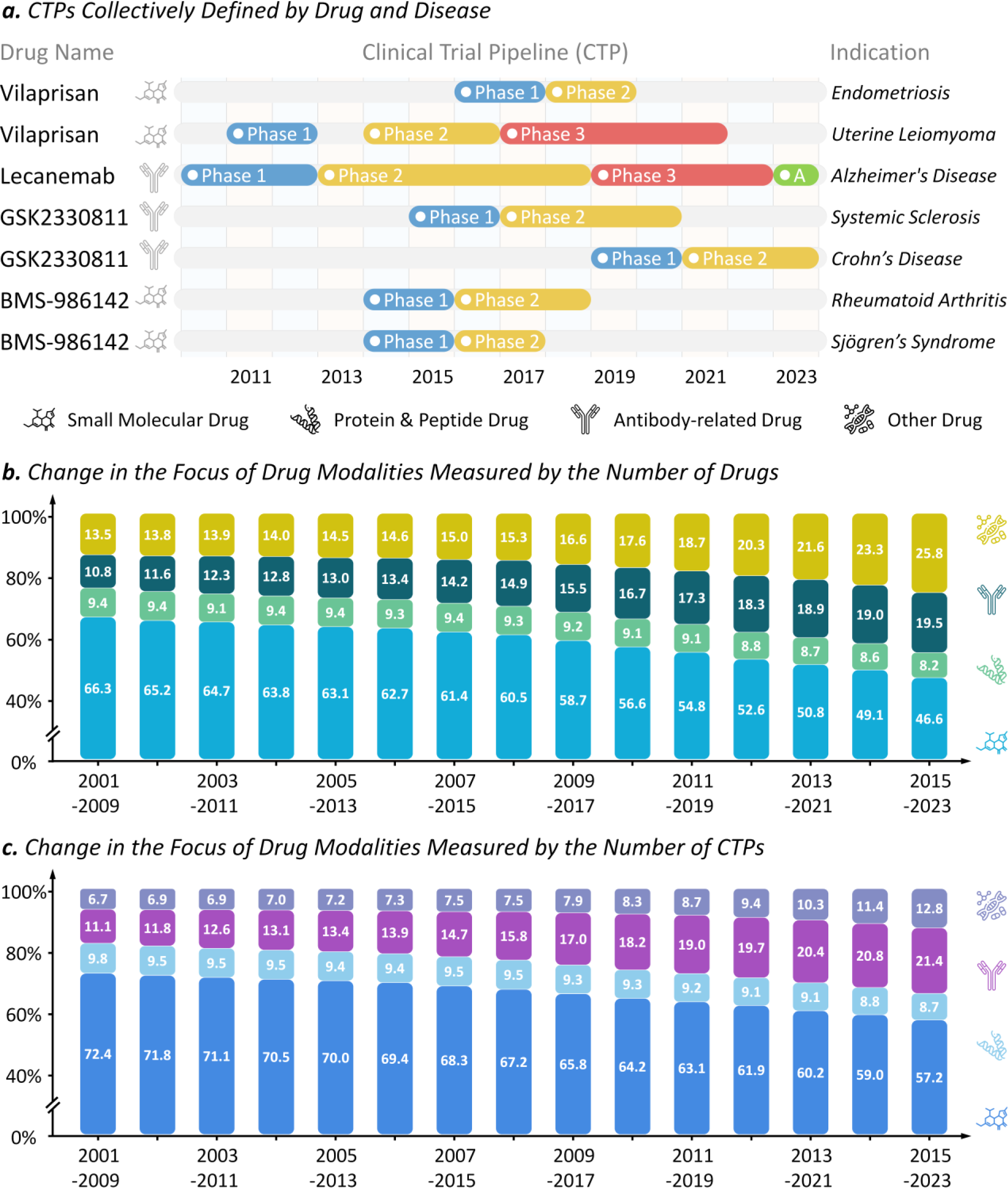
Definition of *clinical trial pipeline* (CTP) and dynamic change in the research focuses of drug modalities. (***a***) the CTPs collectively defined by drug and its corresponding disease. The CTP of a drug for treating one indication was generated by merging all trials of this drug treating the same indication, and those trials of this drug treating other indications were used to generated new CTPs. Taking *vilaprisan* as an example, it was clinically tested for 2 diseases (*endometriosis* and *uterine leiomyoma*), which resulted in 2 distinct CTPs for this drug. (***b***) dynamic shift in the research focus of drug modalities measured by the numbers of clinically tested unique molecular entities in clinical trial. The percentage of small molecular drugs kept declining from 66.3% (at the beginning of 21st century) to 46.6% (now) with observable increase of the shares of antibody-related drugs (from 10.8% to 19.5%) and other drugs (from 13.5% to 25.8%). (***c***) dynamic shifts in research focus of drug modalities measured by the numbers of CTPs. The percentage of CTPs of small molecular drugs kept declining from 72.4% to 57.2% with clear increase of the share of antibody-related drugs (from 11.1% to 21.4%) and others (from 6.7% to 12.8%).

### 2.2 Strategy for Calculating the *Clinical Success Rate* (ClinSR) of Studied Drugs

Before assessing the success rate of clinical trial drugs, a nine-year time-window was adopted in this study to facilitate the definition of clinical success and the calculation of clinical success rate (ClinSR), which gave drug adequate period of time to reach its final fate (12). As a result, a total of fifteen time-windows (from 2001-2009 to 2015-2023, inclusive) were systematically assessed in this study. As shown in **Figure 2*b***, a shift in the research focus on various drug modalities was observed based on assessing the number of unique molecular entities in clinical trial. Particularly, during the past two decades, the percentage of small molecular drugs kept declining from 66.3% (at the beginning of 21st century) to 46.6% (till now) with observable increases of the shares of antibody-related drugs (from 10.8% to 19.5%) and other drugs (from 13.5% to 25.8%, especially RNA-based therapies, cell therapies, gene therapies, *etc.*). Moreover, as illustrated in **Figure 2*c***, a shift in the research focus on various drug modalities was also observed based on assessing the number of CTPs. Particularly, in the past two decades, the percentage of CTPs of small molecular drugs kept declining from 72.4% (early 21st century) to 57.2% (till now) with clear increases of the shares of antibody-related drugs (from 11.1% to 21.4%) and others (from 6.7% to 12.8%).

#### 2.2.1 Describing Progression of Clinical Trial Pipeline (CTP)

To describe the progression of any studied CTP within a time-window, it is critical to know how drug’s clinical status was changed. There were three clinical statuses (Phase 1, Phase 2 and Phase 3) that could be changed in a CTP. Taking the Phase 1 as an example, if it successfully progressed to a higher status (Phase 2, Phase 3 or Approval) in a studied time-window, the progression under Phase 1 was considered as “***Success***” in this study; if it was reported to be clinically discontinued (or terminated) or with no new trials conducted for over two years (13) in a studied time-window, the progression under Phase 1 was regarded as “***Failure***”; otherwise, the progression under Phase 1 was defined as “***Ongoing***”. Similar methodology could be used to describe the progressions of Phase 2 and Phase 3, which could also be categorized into *Success*, *Failure* and *Ongoing*.

#### 2.2.2 Evaluating the Clinical Success Rate (ClinSR) of Drugs

To systematically assess the *clinical success rate* (ClinSR) of drugs within studied time-window (*t*_*begin*_, *t*_*end*_), four key measurements should be calculated, which included: *P*1*SR*(*t*_*begin*_, *t*_*end*_), *P*2*SR*(*t*_*begin*_, *t*_*end*_), *P*3*SR*(*t*_*begin*_, *t*_*end*_), and *OSR*(*t*_*begin*_, *t*_*end*_). Taking the *P*1*SR*(*t*_*begin*_, *t*_*end*_) as an example, 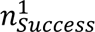 (*t*_*begin*_, *t*_*end*_) indicated the total number of ***Success*** Phase 1 progressions within the time-window, while 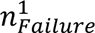(*t*_*begin*_, *t*_*end*_) denoted the total number of ***Failure*** Phase 1 progressions in the same time-window. Therefore, the *P*1*SR*(*t*_*begin*_, *t*_*end*_) could be calculated to represent the success rate for Phase 1 using the following equation:

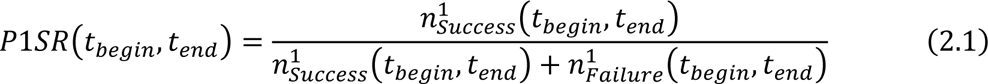

Similarly, the success rates for both Phase 2 and Phase 3 could also be computed. Apart from the three key measurements for assessing ***phase success rate***, *OSR*(*t*_*begin*_, *t*_*end*_) was adopted in this study to denote the ***overall success rate*** from Phase 1 to Approval, which could be calculated by multiplying three *phase success rates P*1*SR*, *P*2*SR*, and *P*3*SR* using the following equation:

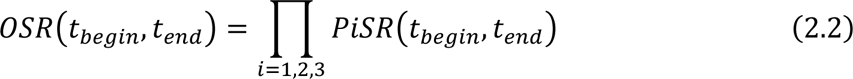

## 3. Results and Discussion

### 3.1 Assessing the Reliability and Comprehensiveness of Collected Data

A landmark study reporting the *clinical success rates* (ClinSRs) of drugs within the time-window of 2003-2011 had been published on *Nature Biotechnology* (2), which had attracted interest from broad research communities. Although the knowledgebase used in that study (*BioMedTracker*, a commercial database) for information collection was different from the one adopted in this work (*Clinicaltrials.gov*), it is of great interests to reproduce the analyses of landmark study using our collected data to see whether there is any bias. As provided in **Table 1**, for all diseases, the *overall success rate* (OSR) calculated by this study for the time-window of 2003-2011 equaled to 10.0%, which was highly consistent with that (10.4%) of landmark study (2), and their *relative difference* (RD) was really small (−3.8%). RD equaled to the *actual difference* (AD) between the values of this work and the landmark study divided by that of landmark study. Moreover, the RDs of P1SR, P2SR and P3SR between this work and the landmark study (described in **Table 1**) were also very small (1.9%, −4.0% and −2.4%, respectively). Thus, these results above showed the reliability of our analytical protocol, and no significant bias was observed in our collected data.

**Table 1.** Comparing the *clinical success rates* (ClinSRs) within the time-window of 2003-2011 between this work and a published landmark study by Hay *et al* (2). The definition of eight disease classes in Hay’s study (2) was directly used by this work to facilitate this comparison. *RD*: *relative difference*, dividing the *actual difference* (AD) between this study and Hay’s study by Hay’s study. The absolute values of all RDs are ≤5%.

| Disease class | OSR<br>(overall success rate) |  |  | P1SR<br>(Phase 1 success rate) |  |  | P2SR<br>(Phase 2 success rate) |  |  | P3SR<br>(Phase 3 success rate) |  |  |
| --- | --- | --- | --- | --- | --- | --- | --- | --- | --- | --- | --- | --- |
|  | This Study | Hay’s Study | RD | This Study | Hay’s Study | RD | This Study | Hay’s Study | RD | This Study | Hay’s Study | RD |
| All Diseases | 10.0% | 10.4% | **-3.8%** | 65.7% | 64.5% | **1.9%** | 31.1% | 32.4% | **-4.0%** | 48.8% | 50.0% | **-2.4%** |
| 1. Infection | 16.1% | 16.7% | **-3.6%** | 64.8% | 65.8% | **-1.5%** | 44.6% | 45.9% | **-2.8%** | 55.7% | 55.4% | **0.5%** |
| 2. Oncology | 6.8% | 6.7% | **1.5%** | 65.2% | 63.9% | **2.0%** | 26.9% | 28.3% | **-4.9%** | 38.6% | 36.9% | **4.6%** |
| 3. Autoimmune | 12.8% | 12.7% | **0.8%** | 71.0% | 68.0% | **4.4%** | 32.5% | 34.0% | **-4.4%** | 55.6% | 54.9% | **1.3%** |
| 4. Endocrine | 11.2% | 11.6% | **-3.4%** | 57.5% | 58.3% | **-1.4%** | 34.0% | 33.8% | **0.6%** | 57.5% | 58.6% | **-1.9%** |
| 5. Neurology | 9.3% | 9.4% | **-1.1%** | 65.3% | 62.4% | **4.6%** | 29.0% | 30.2% | **-4.0%** | 48.9% | 49.8% | **-1.8%** |
| 6. Cardiovascular | 7.4% | 7.1% | **4.2%** | 60.0% | 60.6% | **-1.0%** | 27.6% | 26.3% | **4.9%** | 44.4% | 44.6% | **-0.4%** |
| 7. Respiratory | 10.7% | 11.1% | **-3.6%** | 68.3% | 66.7% | **2.4%** | 26.2% | 27.5% | **-4.7%** | 60.0% | 60.8% | **-1.3%** |
| 8. Other | 17.3% | 18.2% | **-4.9%** | 72.1% | 72.2% | **-0.1%** | 43.2% | 44.2% | **-2.3%** | 55.7% | 57.1% | **-2.5%** |

Moreover, the landmark study (2) also provided an in-depth analysis on ClinSRs for eight disease classes (infection, oncology, autoimmune, endocrine, neurology, cardiovascular, respiratory, and other). In this study, similar in-depth analysis was conducted, and the drugs collected to this work were classified into the same eight classes of disease as that of the landmark study. As illustrated in **Table 1**, for all eight disease classes, the absolute values of the RDs of OSR, P1SR, P2SR and P3SR between this work and the landmark study (2) were always smaller than 5%, which showed great consistence between ClinSRs of this work and that of landmark study. Because of the huge difference among eight disease classes, it is highly challenging to simultaneously reproduce the ClinSRs for eight disease classes. Therefore, the successful reproduction of ClinSRs of previous study further indicated that our collected data were highly reliable for ClinSR assessment.

To ensure the comprehensiveness of our collected data, the quality of both approved and clinical trial drugs were assessed. **Figure 1** gave the numbers of approved *New Drug Applications* (NDA, colored in blue) and *Biologics License Applications* (BLA, colored in pink) collected to this study. The numbers in **Figure 1** were identical to the ones reported on the official website of U.S. FDA and a series of ‘annual FDA approval’ papers published on *Nature Reviews Drug Discovery* (14), which guaranteed the comprehensiveness of the approved drugs collected to this study. Moreover, comparing with other established databases, *ClinicalTrials.gov* had long been considered as one of the most influential sources of clinical trial drugs and clinical testing information, which had rapidly expanded since 2007 due to the official supports from U.S. FDA (*2007 FDA Amendments Act* required all clinical trials to be registered to *Clinicaltrials.gov*). Therefore, *ClinicalTrials.gov* was adopted as the only source for collecting clinical trial drug data, which ensured the reliability of clinical data and maintained the high criteria of data inclusion across different years.

### 3.2 Measuring the Dynamic Change of ClinSRs for All Studied Drugs

With the dramatic investment increase and continuous technological advance during the past two decades (15), researchers are highly curious about how *clinical success rate* (ClinSR) is affected over time. Herein, the dynamic ClinSRs of 15 time-windows from the beginning of 21st century to now were therefore systematically assessed. As illustrated in **Figure 3**, the *phase success rates* (PSRs) of P1SR, P2SR and P3SR were described using bars in blue, yellow and red, respectively, and the dark line with dots indicated the dynamic variation in *overall success rate* (OSR). It was clear that the OSRs had been declining over time and remained stable around 5% in recent years. Comparing with the OSR (10.4%) reported in a previous landmark study (2), the OSRs of recent years were cut by half. In other words, despite the tremendous efforts made to almost every step of drug development (16), the progress of current drug discovery was still in a dilemma.

**Figure 3.**
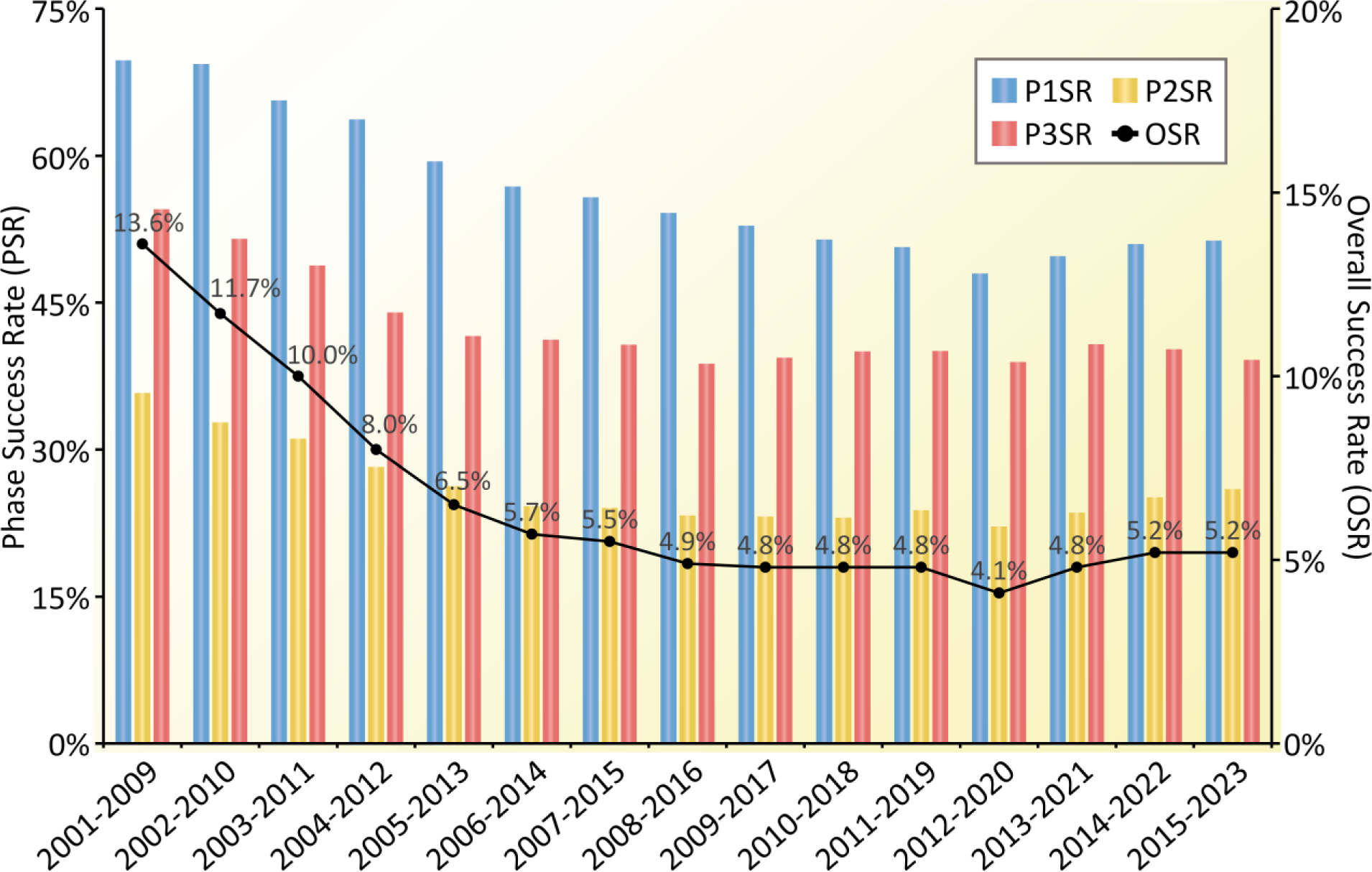
The dynamic *clinical success rate* (ClinSR) evaluated based on all CTPs collected for this study. A nine-year time-window was adopted to facilitate the assessments of ClinSRs, which offered a drug adequate period of time to reach its final fate (12), and a total of 15 time-windows (from 2001-2009 to 2015-2023, inclusive) were systematically assessed. The *phase success rates* (PSRs) of P1SR, P2SR and P3SR were described using bars in blue, yellow and red, respectively. The dark line with dots indicated the dynamic change in *overall success rate* (OSR). It was clear that the OSR had been declining over time and remained stable around 5% in recent years. P1SR: Phase 1 success rate; P2SR: Phase 2 success rate; P3SR: Phase 3 success rate.

An in-depth investigation of **Figure 3** showed that the P2SR (yellow) of every time-window was consistently lower than P1SR (blue) and P3SR (red), which indicated that efficacy (mainly tested in Phase 2) remained one of the largest obstacles in current drug development (17). Furthermore, P1SR (blue) was found to continuously decline from ∼70% to ∼50% during the past two decades. As reported, the objectives of current Phase 1 studies were gradually expanded to evaluate some part of pharmacokinetics/pharmacodynamics and efficacy besides the previous safety evaluation (18), and the so-called ‘quick-kill’ strategy rapidly adopted in pharmaceutical companies brought up more drug candidates to terminate the inferior ones in an earlier stage, especially Phase 1 (19). All these important factors collectively contributed to the persistent decline of P1SR.

It was also shown in **Figure 3** that P3SR (red) had gradually declined since the beginning of this century, and in contrast to both P1SR and P2SR, the P3SR demonstrated further decline in recent time-windows (from 2013-2021 to 2015-2023). It was reported that comparing with the protocol design of Phase 3 in 2001-2005, the complexity of that in 2011-2015 had increased by 70% (20). Increased complexities further led to longer cycle time, higher numbers of protocol amendments, or lower patient recruitment/retention rate (20), which greatly contributed to the clear decline of P3SR in the first eight time-windows of **Figure 3**. Moreover, the further decline of P3SRs in the recent three time-windows of **Figure 3** mainly came from the dramatic decrease of P3SR in some major disease classes, such as *infectious/parasitic disease*, *metabolic disease*, *circulatory system disease*, and so on. Taking the *infectious/parasitic disease* as example, tremendous clinical trials for COVID-19 were tested, and the majority of the Phase 3 clinical trials were reported to end in failure (21), which contributed to the decline of P3SR in recent years. The impact of COVID-19 related clinical trials on ClinSR will be further discussed in the following section.

### 3.3 Diverse and Dynamic ClinSRs Measured based on Disease Classes

In addition to the ClinSR for all CTPs, it was of great interests to further evaluate the ClinSR for CTPs of specific disease class. As shown in **Table 2**, the *overall success rates* (OSRs) of fourteen disease classes (defined by the WHO ICD-11) across fifteen time-windows were systematically offered. Taking the latest time-window 2015-2023 as an example, there was substantial variation in the OSRs (from 2.9% to 17.2%) of different classes of disease, which reminded us to perform further assessment on disease-specific ClinSRs. Therefore, a review on the data collected to this study was conducted, which identified three disease classes that covered the highest numbers of CTPs: *oncologic diseases*, *neurological diseases*, and *infectious/parasitic diseases*. These three classes had long been regarded as three of the most common research domains in both academia and industry (22,23), which required an in-depth analysis in the following sections.

**Table 2.** The *overall success rate* (OSR) for all CTPs collected for this study and the CTPs of specific disease class (a total of 14 classes defined by WHO ICD-11) calculated across fifteen nine-year time-windows. All: all diseases; INFEC: Infectious/parasitic disease; CACER: Oncology; BLOOD: Blood/blood-forming organs disease; IMMUN: Immune system disease; METAB: Endocrine, nutritional or metabolic disease; NEURO: Neurology; VISAL: Visual system disease; CIRCU: Circulatory system disease; RESPR: Respiratory system disease; DIGST: Digestive system disease; SKINS: Skin disease; MUSKE: Musculoskeletal system/connective tissue disease; GENIT: Genitourinary and sexual related disease; OTHER: Other disease.

| Disease Class | 2001<br>-2009 | 2002<br>-2010 | 2003<br>-2011 | 2004<br>-2012 | 2005<br>-2013 | 2006<br>-2014 | 2007<br>-2015 | 2008<br>-2016 | 2009<br>-2017 | 2010<br>-2018 | 2011<br>-2019 | 2012<br>-2020 | 2013<br>-2021 | 2014<br>-2022 | 2015<br>-2023 |
| --- | --- | --- | --- | --- | --- | --- | --- | --- | --- | --- | --- | --- | --- | --- | --- |
| All | 13.6% | 11.7% | 10.0% | 7.9% | 6.5% | 5.7% | 5.5% | 4.9% | 4.8% | 4.8% | 4.8% | 4.1% | 4.8% | 5.2% | 5.2% |
| 01 INFEC | 21.7% | 20.1% | 16.1% | 12.3% | 8.0% | 7.5% | 7.9% | 6.7% | 5.5% | 4.9% | 4.4% | 3.2% | 5.1% | 5.0% | 3.1% |
| 02 CACER | 8.1% | 7.1% | 6.7% | 5.6% | 5.1% | 4.1% | 3.9% | 3.7% | 3.7% | 3.7% | 3.7% | 3.2% | 3.5% | 3.6% | 3.9% |
| 03 BLOOD | 53.1% | 47.3% | 35.4% | 34.6% | 28.8% | 24.3% | 22.5% | 19.9% | 22.0% | 19.6% | 21.0% | 19.7% | 17.7% | 17.3% | 17.2% |
| 04 IMMUN | 23.8% | 32.7% | 22.6% | 18.7% | 13.4% | 11.5% | 8.5% | 6.2% | 7.5% | 7.4% | 6.4% | 5.1% | 8.0% | 9.4% | 10.0% |
| 05 METAB | 22.0% | 16.5% | 11.2% | 9.2% | 7.5% | 7.3% | 7.4% | 7.0% | 7.4% | 8.2% | 8.5% | 7.8% | 9.1% | 9.1% | 9.6% |
| 06 NEURO | 14.8% | 11.7% | 9.1% | 6.1% | 4.6% | 3.8% | 3.5% | 3.4% | 3.6% | 3.6% | 4.5% | 4.5% | 4.7% | 5.8% | 6.3% |
| 07 VISAL | 20.0% | 17.9% | 15.9% | 14.3% | 11.3% | 10.7% | 8.0% | 8.4% | 8.2% | 6.5% | 5.1% | 3.9% | 4.8% | 6.2% | 6.5% |
| 08 CIRCU | 12.6% | 9.4% | 7.4% | 6.4% | 6.2% | 6.6% | 7.1% | 5.8% | 5.7% | 5.6% | 4.9% | 4.0% | 4.4% | 3.8% | 2.9% |
| 09 RESPR | 13.6% | 10.4% | 10.7% | 8.3% | 6.6% | 6.7% | 6.5% | 5.7% | 5.0% | 4.9% | 4.8% | 3.8% | 5.1% | 5.2% | 5.1% |
| 10 DIGST | 11.1% | 9.5% | 9.7% | 8.4% | 8.3% | 5.6% | 5.5% | 4.1% | 4.6% | 5.9% | 5.9% | 4.5% | 5.4% | 6.6% | 6.4% |
| 11 SKINS | 9.7% | 12.9% | 9.0% | 4.8% | 3.2% | 4.5% | 6.0% | 5.4% | 5.8% | 7.0% | 6.7% | 6.9% | 8.3% | 9.6% | 11.2% |
| 12 MUSKE | 27.7% | 21.2% | 18.8% | 16.1% | 13.4% | 11.3% | 9.9% | 8.7% | 7.8% | 7.3% | 7.9% | 6.8% | 7.3% | 7.8% | 9.3% |
| 13 GENIT | 24.0% | 17.0% | 10.2% | 6.9% | 9.1% | 9.3% | 8.3% | 8.3% | 7.2% | 8.5% | 10.5% | 6.9% | 5.2% | 4.9% | 5.4% |
| 14 OTHER | 15.1% | 10.8% | 14.3% | 10.0% | 8.6% | 6.9% | 6.9% | 4.2% | 3.8% | 2.8% | 2.6% | 3.1% | 4.1% | 3.9% | 4.4% |

#### 3.3.1 Assessing the ClinSRs for Drugs Treating Oncologic Disease

The dynamic ClinSRs evaluated based on the CTPs of *oncologic diseases* collected for this study were explicitly described in **Supplementary Figure S1**. As shown, the OSRs had been declining over time; since the time-window of 2006-2014, the OSRs kept below 5% with small fluctuation among recent time-windows. Such results were consistent with a recent study (24) reporting that the success rate in anticancer drug developments was less than 5%, and the potential contributors to such low success rate might include limited understanding of cancer biology, poorly predictive preclinical models, and heterogeneity among patients (25,26). On the one hand, P2SR was found consistently lower than P1SR and P3SR, which indicated that Phase 2 remained the largest driver of the clinical failure for anticancer drug development (27). On the other hand, in contrast to the clear increase of P3SR from 37.5% to 55.1% (as provided in **Supplementary Figure S1**), P1SRs dramatically declined from 67.8% to 39.1%. Such declines in P1SRs pointed out increasing risks in the early clinical development of innovative targeted drug and immunotherapy for cancer (24), which recently prompted the U.S. FDA *Oncology Center of Excellence* (OCE) to launch ‘*Project Optimus*’ focusing on the dose optimization for Phase 1 trial of anticancer therapy discovery (28). In addition, the increase of P3SR accompanied by decline of P1SR identified in this study might indicate that the early clinical evaluation of current pharmaceutical industry became increasingly thorough, which might help to prevent the costly late-stage (especially Phase 3) failure (13).

According to the clinical trial data collected, anticancer drugs consisted of the largest proportion among other disease classes, and it was therefore essential to investigate the impacts of oncologic therapies on the ClinSR of all CTPs. In this study, the comparison of ClinSRs between oncologic (yellow) and non-oncologic (blue) CTPs was provided in **Figure 4*a***. As illustrated, the OSRs of the CTPs of the anticancer drugs (oncologic) were consistently lower than that of non-anticancer (non-oncologic) ones. Particularly, although P1SRs of oncologic and non-oncologic CTPs were comparable in the beginning of 21st century, the oncologic P1SRs showed continuous decline in recent years, which was different from the trend of slight increase of non-oncologic P1SRs; when it came to P2SR, the evolving trends of oncologic and non-oncologic CTPs were almost identical with the non-oncologic P2SRs consistently higher than oncologic ones; in contrast to the decline trend of non-oncologic P3SRs, the oncologic P3SRs persistently increased. All in all, significant impacts of oncologic therapies on the ClinSR of all CTPs were observed in this study.

**Figure 4.**
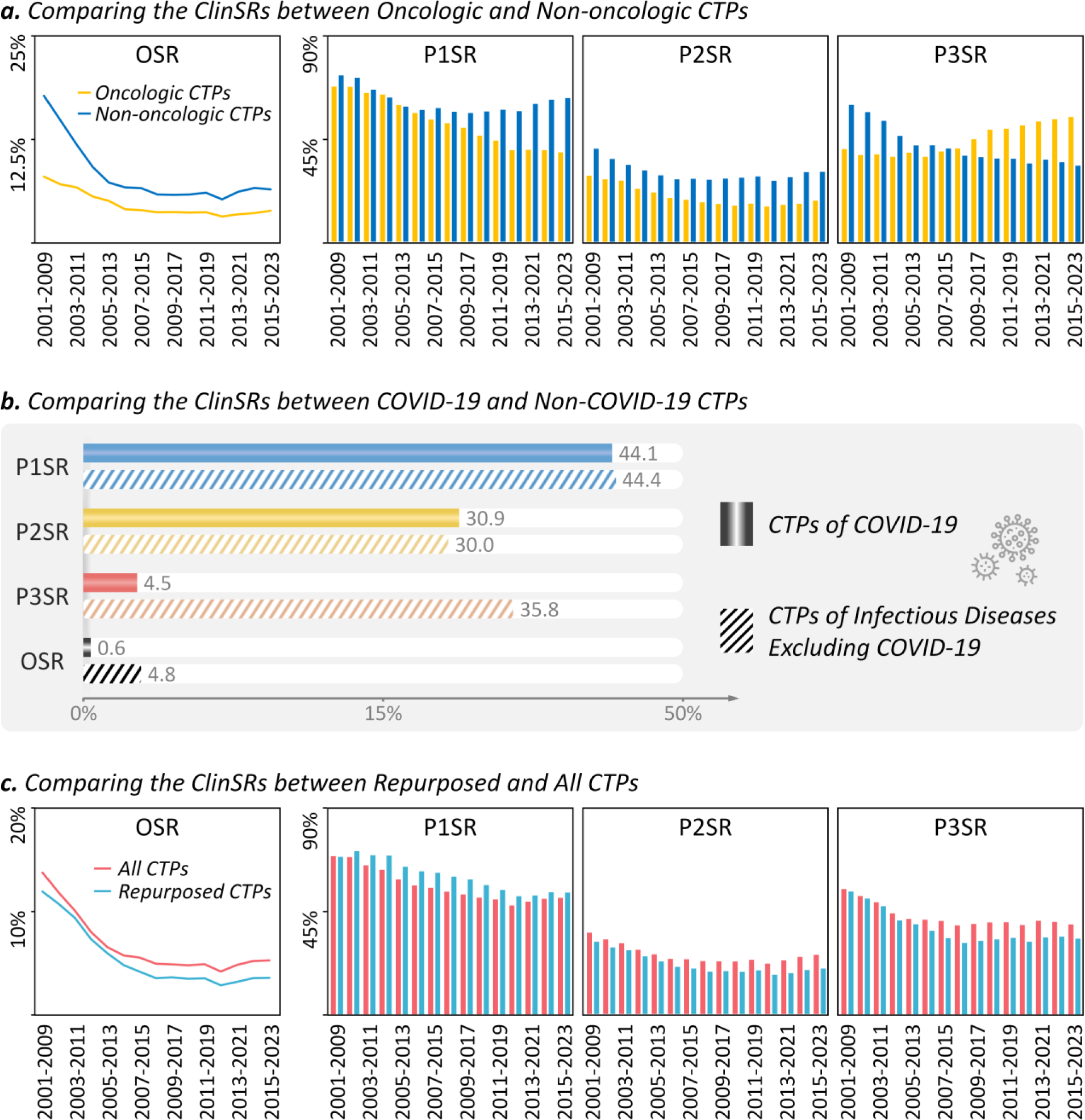
Comparing the ClinSRs between the CTP groups of different disease classes or clinical discovery strategies. (***a***) comparing the ClinSRs between oncologic and non-oncologic CTPs. (***b***) comparing the ClinSRs between the CTPs of COVID-19 and that of infectious disease excluding COVID-19. (***c***) comparing the ClinSRs between repurposing CTPs and all collected CTPs. CTP: clinical trial pipeline; ClinSR: clinical success rate; P1SR: Phase 1 success rate; P2SR: Phase 2 success rate; P3SR: Phase 3 success rate; OSR: overall success rate.

#### 3.3.2 Assessing the ClinSRs for Drugs Treating Neurological Disease

The dynamic ClinSRs evaluated using the CTPs of *neurological diseases* collected for this study were explicitly described in **Supplementary Figure S2**. As shown, the OSRs had been declining in the early 21st century by hitting the bottom with an extremely low OSR of 3.4% in 2008-2016, and then experienced a slow but clear increase in recent years. As reported, such extremely low OSR of neurological diseases primarily came from the difficulty in crossing *blood-brain barrier*, notoriously unpredictive animal models, and poor understanding of complex CNS condition (29). To deal with such problems, new technologies/models (such as targeted protein degradation, gut-microbiota interventions, and stem cell therapy) had been adopted in the past decade (30), which might substantially contribute to the steady elevation in the OSR of neurological disease in recent years (shown in **Supplementary Figure S2**). Moreover, the evolution of *phase success rate* was also described in **Supplementary Figure S2**. Comparing with P1SRs and P2SRs, there was clear elevation in recent P3SRs, which contributed the most to the recent elevation of OSR.

#### 3.3.3 Assessing the ClinSRs for Drugs Treating Infectious/parasitic Disease

The dynamic ClinSRs evaluated based on the CTPs of *infectious diseases* collected for this study were explicitly described in **Supplementary Figure S3**. As shown, the OSRs had been declining over time, and hit the bottom with a very low OSR of 3.1% in the latest time-window 2015-2023. At the beginning of this century, the OSR of infectious disease was more than two times as many as that of oncology, while its OSR in recent years became comparable to that of oncology, which documented a dramatic decline in its ClinSR. As reported, the development of anti-infective drug had changed its pivot from non-host targets to host targets, which led to increasing development difficulty and might therefore result in the dramatic decline of clinical success (25).

The drugs/candidates for treating COVID-19 had been frequently tested in clinical trial in recent years, which consisted of a large proportion of anti-infective drugs, and it was therefore essential to investigate the impact of COVID-19 therapies on the ClinSR of all anti-infective drugs. In this study, the comparison of ClinSRs between the CTPs of COVID-19 and that of infectious diseases excluding COVID-19 was conducted, and the results were described in **Figure 4*b***. As illustrated, there was no significant difference in P1SRs and P2SRs between the studied two groups of CTPs. However, dramatic variation was observed in P3SR which provided a significantly lower rate of success (4.5%) for COVID-19 CTPs than that (35.8%) of non-COVID-19 CTPs. Moreover, such a low P3SR further resulted in a low OSR (0.6%) of COVID-19 CTPs comparing with that (4.8%) of non-COVID-19 CTPs. Recent study (21) found that most COVID CTPs successfully entered in Phase 3, while the vast majority were reported to end in failure. In other words, although there were drugs successfully approved for COVID-19 in significantly short time, it was apparent that these successes came at the high cost of a huge number of clinical trial failures.

Besides those three disease classes discussed above, the dynamic ClinSRs assessed based on the CTPs of eleven additional classes of disease (such as: *circulatory system disease*) defined by the WHO ICD-11 were explicitly shown in **Supplementary Figure S4**-**S14**. The detailed values of the calculated P1SRs, P2SRs, P3SRs were also provided in **Supplementary Table S2**-**S4**.

#### 3.3.4 Similarity among Disease Classes Identified Based on Their ClinSRs

To reveal the similarity among diseases in their ClinSRs across fifteen time-windows, the cluster analyses based on OSRs, P1SRs, P2SRs and P3SRs were carefully conducted, and corresponding results were showed in **Figure 5**, **Supplementary Figure S15**, **Supplementary Figure S16** and **Supplementary Figure S17**, respectively. Particularly, various disease classes were *first* ranked based on the ClinSRs across fifteen time-windows, and complete linkage hierarchical clustering was *then* calculated using the ranking results based on *Euclidean* distance. Taking the clustering based on OSRs (shown in **Figure 5**) as an example, two distinct clustering groups were identified with six disease classes (BLOOD, MUSKE, IMMUN, METAB, GENIT & VISAL) at the bottom and eight classes (CACER, CIRCU, NEURO, DIGST, RESPR, INFEC, SKINS & OTHER) on the top. As a typical disease of the bottom group, blood/blood-forming organs disease (BLOOD) was found with consistently higher OSRs than other diseases, which made it similar to other two classes of disease: immune system disease (IMMUN), musculoskeletal system/connective tissue disease (MUSKE). Moreover, oncology (CACER) and circulatory system disease (CIRCU) were found to be typical disease classes of the top group, which provided consistently the lowest OSRs across fifteen time-windows comparing with other disease classes.

**Figure 5.**
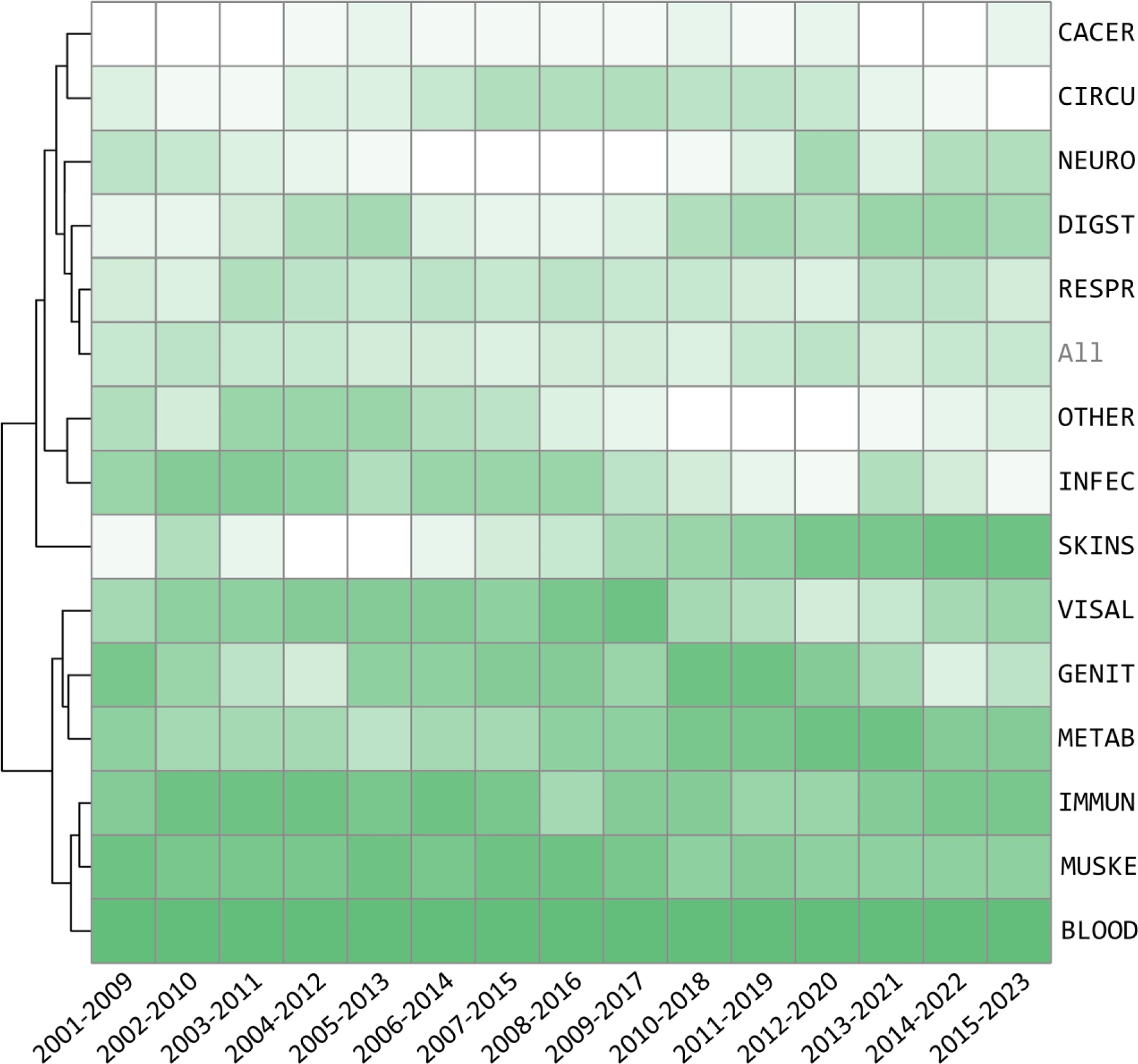
Hierarchical clustering of different disease classes based on their *overall success rates* (OSRs) across 15 time-windows. The darker the green color, the higher the success rate for drug development in the corresponding disease and time-window. INFEC: infectious/parasitic disease; BLOOD: blood/blood-forming organs disease; CACER: oncology; CIRCU: circulatory system disease; DIGST: digestive system disease; SKINS: skin disease; METAB: endocrine, nutritional or metabolic disease; RESPR: respiratory system disease; MUSKE: musculoskeletal system and connective tissue disease; GENIT: genitourinary and sexual related disease; NEURO: neurology; VISAL: visual system disease; IMMUN: immune system disease; OTHER: other disease.

#### 3.3.5 Assessing the ClinSRs for Repurposed Drugs

Drug repurposing was a strategy to discover new indication for drug beyond its initial indication (31). Given its characteristic of the less risk in safety, more rapid return on investment, and lower average cost after failure, the enthusiasm for drug repurposing was growing (32). An appreciable number of pharmaceutical researchers hold an optimistic attitude that drug repurposing was more likely to be successful than traditional way of drug development (33). However, there was a lack of the systematic and quantitative analyses on such point of view.

In this study, a comparison of ClinSRs between repurposed CTPs and all CTPs was provided in **Figure 4*c***. As illustrated, the OSRs of the repurposed CTPs were consistently lower than that of all CTPs collected to this study, which was, from the perspective of ClinSRs at least, contrary to the ‘optimistic attitude’ on the success of repurposed drug. Particularly, the P1SRs of repurposed CTPs remained higher than that for all CTPs across all fifteen time-windows, which was readily understandable since most repurposed drugs had been previously assessed for safety. Meanwhile, both P2SRs and P3SRs of repurposed CTPs were illustrated in **Figure 4*c*** to be consistently lower than that for all CTPs. The relatively low cost of trial-and-error in drug repurposing had promoted a large number of trials to rush into the clinical test without clear understanding of the underlying disease biology and target mechanism (25,34), which might substantially contribute to the failure of many repurposed drugs and in turn the relatively low overall success rates. The analyses above also reminded us about the low ClinSRs identified in previous section when developing COVID-19 drugs, most of which were repurposed ones (35). All in all, although drug repurposing is well-intentioned and attractive, available evidence suggests that cautions should be taken.

### 3.4 Diverse and Dynamic ClinSRs Measured based on Drug Modalities

Drug modality had also been considered as one of the risk contributors to the success rate of drug development (36). Small molecular drug (SMD) had long been the dominant modality and newer ones (such as: antiboday-related drug) had also been added to the drug development toolbox (37). In this study, the analyses on four types of major drug modality, including *small molecular drug* (SMD), antibody-related drug (ARD), protein & peptide drug (PPD) and other drug (OTH), were conducted, and their ClinSRs were systematically provided in **Table 3**, and separately described in **Supplementary Figure S18-21**. As illustrated in **Table 3**, the OSRs of ARD were consistently higher than that of other modalities in recent decade, while the OSRs of PPD at the beginning of the 21st century surpassed that of the others. The OSRs of OTH (a mixture of very diverse types of drug, such as: RNA therapy and cell therapy) remained the lowest across fifteen time-windows. As the most well-established drug modality, three factors of SMD were considered as the primary reason leading to its failure, including poor physicochemical property, unmeaningful efficacy of the chosen targets and constant turmoil of strategy changes with pharmaceutical companies (37). With the increasing elucidation of the molecular mechanism underlying the disease pathogenesis, significant growth potential of antibody-related drugs was also highly anticipated (38). Because of the unique advantages of different drug types, current pharmaceutical industry tended to adopt a broad mixture of drug modalities for disease treatment (37).

**Table 3.** The *clinical success rates* (ClinSR, assess both *overall success rate* and *phase success rate*) for various drug modalities in the 21st century calculated within nine-year time-window. SMD: small molecular drug; ARD: antibody-related drug; PPD: protein & peptide drug; OTH: other drug; OSR: overall success rate; P1SR: Phase 1 success rate; P2SR: Phase 2 success rate; P3SR: Phase 3 success rate.

| Drug Modality |  | 2001<br>-2009 | 2002<br>-2010 | 2003<br>-2011 | 2004<br>-2012 | 2005<br>-2013 | 2006<br>-2014 | 2007<br>-2015 | 2008<br>-2016 | 2009<br>-2017 | 2010<br>-2018 | 2011<br>-2019 | 2012<br>-2020 | 2013<br>-2021 | 2014<br>-2022 | 2015<br>-2023 |
| --- | --- | --- | --- | --- | --- | --- | --- | --- | --- | --- | --- | --- | --- | --- | --- | --- |
| OSR | SMD | 12.3% | 10.1% | 8.6% | 6.7% | 5.8% | 4.8% | 4.6% | 4.1% | 3.9% | 4.0% | 4.0% | 3.5% | 3.9% | 4.1% | 4.1% |
|  | ARD | 21.5% | 20.7% | 19.7% | 17.8% | 13.9% | 12.1% | 11.6% | 10.3% | 10.0% | 9.3% | 9.9% | 7.8% | 9.6% | 11.0% | 11.3% |
|  | PPD | 24.6% | 24.3% | 19.1% | 16.3% | 12.0% | 12.3% | 10.7% | 8.8% | 8.4% | 7.5% | 6.4% | 4.8% | 5.0% | 4.9% | 5.3% |
|  | OTH | 1.3% | 5.0% | 3.3% | 2.2% | 1.6% | 1.9% | 2.0% | 2.1% | 2.5% | 2.3% | 2.5% | 2.2% | 3.1% | 3.4% | 3.7% |
| P1SR | SMD | 71.3% | 70.9% | 67.1% | 65.6% | 62.7% | 60.1% | 58.6% | 57.9% | 56.7% | 55.1% | 53.8% | 52.4% | 53.4% | 54.9% | 53.7% |
|  | ARD | 63.3% | 65.2% | 65.9% | 64.1% | 59.1% | 57.2% | 56.0% | 51.7% | 50.6% | 48.4% | 48.6% | 44.3% | 47.9% | 50.0% | 52.7% |
|  | PPD | 69.6% | 72.1% | 64.4% | 64.8% | 59.6% | 58.1% | 58.3% | 56.3% | 54.0% | 54.7% | 53.0% | 49.0% | 49.4% | 51.5% | 52.1% |
|  | OTH | 67.1% | 61.6% | 55.3% | 49.7% | 40.3% | 37.4% | 36.9% | 35.9% | 34.8% | 34.7% | 35.3% | 31.8% | 35.5% | 35.6% | 39.2% |
| P2SR | SMD | 32.7% | 29.4% | 27.7% | 25.0% | 23.7% | 21.4% | 21.5% | 20.8% | 20.5% | 20.5% | 21.2% | 20.2% | 21.0% | 22.2% | 23.2% |
|  | ARD | 49.2% | 46.6% | 45.6% | 43.0% | 37.9% | 36.1% | 34.2% | 32.2% | 31.3% | 31.4% | 32.9% | 28.8% | 32.3% | 34.5% | 34.7% |
|  | PPD | 50.0% | 46.9% | 46.0% | 40.4% | 36.9% | 37.1% | 34.3% | 32.0% | 31.7% | 29.9% | 28.5% | 25.1% | 24.3% | 25.4% | 26.8% |
|  | OTH | 31.4% | 34.0% | 28.8% | 26.1% | 22.9% | 21.1% | 20.5% | 21.3% | 23.2% | 22.5% | 23.4% | 21.6% | 24.5% | 26.4% | 26.5% |
| P3SR | SMD | 52.8% | 48.6% | 46.4% | 40.9% | 38.9% | 37.6% | 36.5% | 33.7% | 33.5% | 34.9% | 35.3% | 33.5% | 34.6% | 33.8% | 32.7% |
|  | ARD | 68.9% | 68.0% | 65.5% | 64.5% | 61.8% | 58.7% | 60.4% | 62.0% | 63.3% | 61.5% | 61.9% | 61.6% | 62.3% | 63.4% | 61.5% |
|  | PPD | 70.8% | 72.0% | 64.4% | 62.1% | 54.4% | 57.0% | 53.6% | 48.6% | 48.7% | 46.1% | 42.1% | 38.7% | 41.8% | 37.7% | 37.7% |
|  | OTH | 6.3% | 24.0% | 20.7% | 17.1% | 17.5% | 24.5% | 26.3% | 27.0% | 31.3% | 28.9% | 30.0% | 32.0% | 35.8% | 35.6% | 35.8% |

### 3.5 Construction of Multi-Functional Platform for Reporting ClinSRs

The ClinSRs of drugs were critical for both clinical researcher and pharmaceutical investor when making scientific and economic decisions (4). However, the serious problem of ‘information lag’ of previous studies could not effectively demonstrate the dynamic nature of ClinSR. Furthermore, considering the diverse research interests among researchers, a customized analysis on particular groups of drugs was highly demanded, but no such tool had been available. In this study, a multi-functional online platform, entitled ‘ClinSR.org’, was thus constructed, which enabled a dynamic description of the ClinSR of any drug group of interests. Moreover, to cope with the problem of information lag, ClinSR.org was carefully designed to not only integrate all the data collected to this study, but also promise to continuously update new information for next decade. The unique characteristics of this newly developed online platform were explicitly described as follows.

### An Automated Platform Enabling the Dynamic Description of ClinSRs

As shown in **Figure 6*a***, a process enabling the automated data collection and ClinSR assessment was constructed. *First*, drugs and their corresponding clinical status were automatically collected from *ClinicalTrials.gov* and the U.S. FDA website by quarterly retrieving information using their *Application Programming Interface* (API). *Second*, diverse data affiliated to the newly collected drugs were automatically retrieved by matching with three established databases (WHO ICD-11, DrugBank and TTD). *Third*, all the collected data were carefully reviewed and validated by well-trained pharmacologists and bioinformaticians in our team to guarantee the data quality, and were then integrated into the large pool of data collected in this study. *Finally*, the dynamic change of ClinSRs among time-windows were automatically calculated using the latest collection of drugs, which was then timely updated and systematically visualized on the website of ClinSR.org. Our team promised to persistently update new data to ClinSR.org for the coming decade.

**Figure 6.**
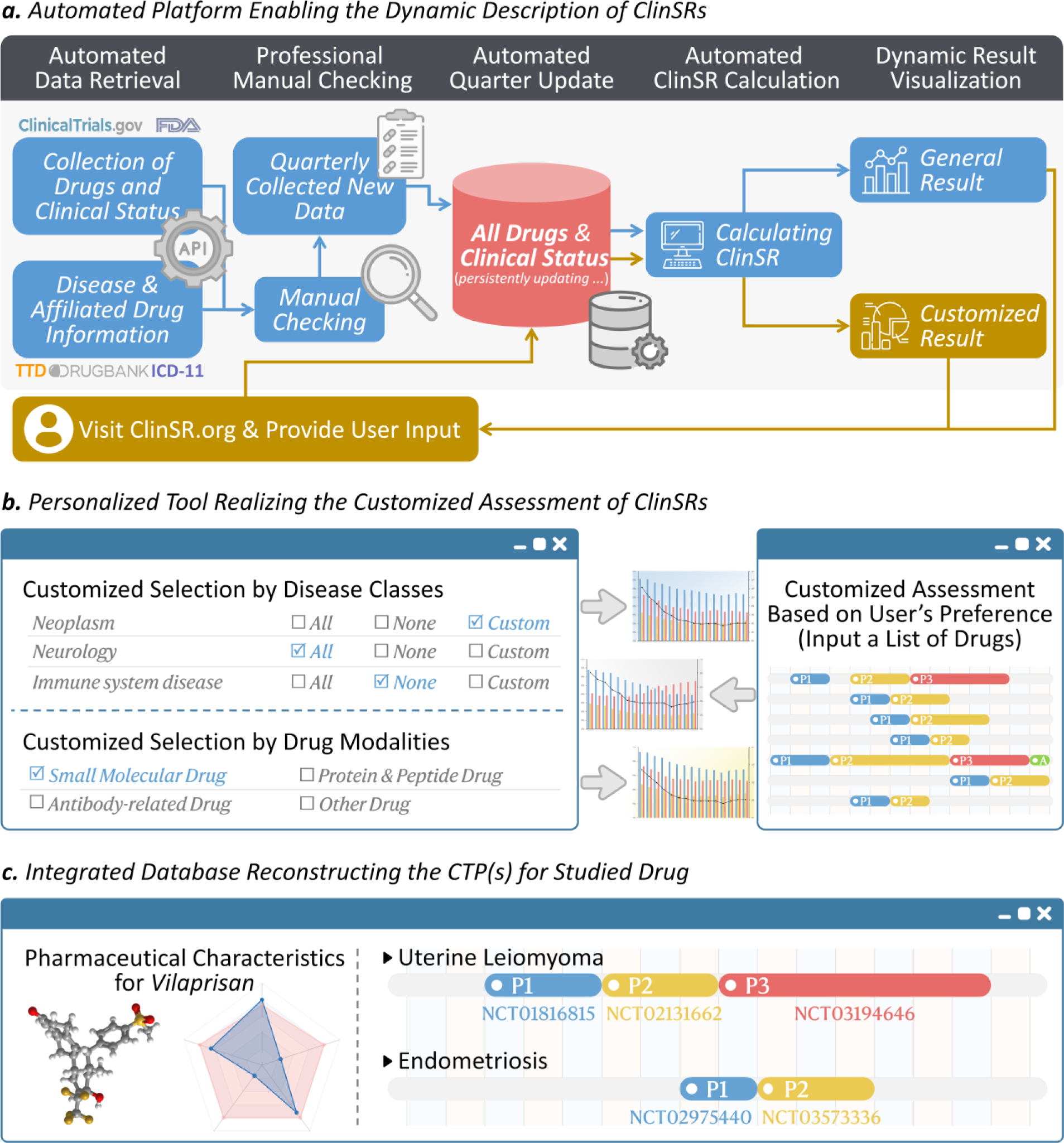
The multi-functional online platform *ClinSR.org* constructed in this study. The unique characteristics of *ClinSR.org* included: (***a***) automated platform enabling the dynamic description of ClinSRs; (***b***) personalized tool realizing the customized assessment of ClinSRs; (***c***) integrated database reconstructing the CTP(s) for studied drug. Our research team promised to persistently update new data and provide the latest ClinSR information to *ClinSR.org* for the coming decade.

### A Personalized Tool Realizing the Customized Assessment of ClinSRs

As illustrated in **Figure 6*b***, a variety of strategies realizing the customized assessment of ClinSR based on the user’s preference were provided in ClinSR.org. Particularly, a user was allowed to assess the ClinSR for a particular class of disease or a specific modality of drug, and also evaluate the joint contribution of multiple disease classes or drug modalities to the success of clinical trial drugs. Moreover, the ClinSR.org enabled the assessment of ClinSR for any drug group of interest. User can first upload a list of drugs (indicated by drug name, TTD drug ID, DrugBank accession, PubChem CID, *etc.*), and the ClinSR of these drugs will then be automatically calculated.

### An Integrated Database Reconstructing the CTP(s) for Studied Drug

Although *ClinicalTrials.gov* offered extensive clinical information on trial drugs, it lacked a clear description of the *clinical trial pipelines* (CTPs) for individual drug. Particularly, the information on *ClinicalTrials.gov* was offered in pieces, each of which focused only on one trial, which asked for the reconstruction of the entire CTP for each drug. As illustrated in **Figure 6*c***, the CTPs were therefore systematically reconstructed for each drug collected to this study, which were explicitly described in the “*Pipeline Identification for a Drug of Distinct Disease*” section of **Material and Methods**. Taking *vilaprisan* (illustrated in **Figure 6*c***) as an example, it had been clinically tested for two disease indications (*endometriosis* and *uterine leiomyoma*). This led to two distinct CTPs for this specific drug, which were systematically described in ClinSR.org to facilitate the decision making for the researchers and investors in the fields of pharmaceutical sciences.

## 4. Conclusion

In this study, a systematic analysis on the dynamic clinical success rate (ClinSR) of drugs in 21st century was first conducted, and a multi-functional platform entitled ‘ClinSR.org’ was developed online (http://ClinSR.idrblab.org/) to realize dynamic description of how ClinSR changes over time, enable automated update of ClinSRs for the coming new decade, and allow the customized evaluation of ClinSR for any drug group of interest. To the best of our knowledge, this study was expected to effectively support the decision making in current drug discovery.

## Data Availability

All data produced are available online at: http://ClinSR.idrblab.org/

## Acknowledgement

Funded by National Natural Science Foundation of China (82373790, 22220102001, 81872798 & U1909208); Natural Science Foundation of Zhejiang Province (LR21H300001); Fundamental Research Fund for Central Universities (2018QNA7023); National Key R&D Program of China (2022YFC3400501); Double Top-Class University (181201*194232101); Key R&D Program of Zhejiang (2020C03010); Westlake Laboratory (Westlake Lab of Life Sciences and Biomedicine); Alibaba-ZJU Joint Research Center of Future Digital Healthcare.

## Authors’ Contribution

F.Z. conceived the idea and designed the entire research; Yi.Z., Y.T.Z., Z.C., S.J.H., Y.H.L., J.B.F, D.H.Z. and X.C.L. collected the data; Yi.Z., Y.T.Z. and H.N.Z. performed the data analyses; Yi.Z., Y.T.Z., Z.C., S.J.H., Yu.Z., Y.Q.Q. and L.Y.H. produced all the figures and tables; Yi.Z. and Y.T.Z. designed and constructed the ClinSR.org website; F.Z. and Y.Z. wrote the manuscript.

