## Supplementary for "Dynamic Clinical Success Rates for Drugs in the 21st Century"

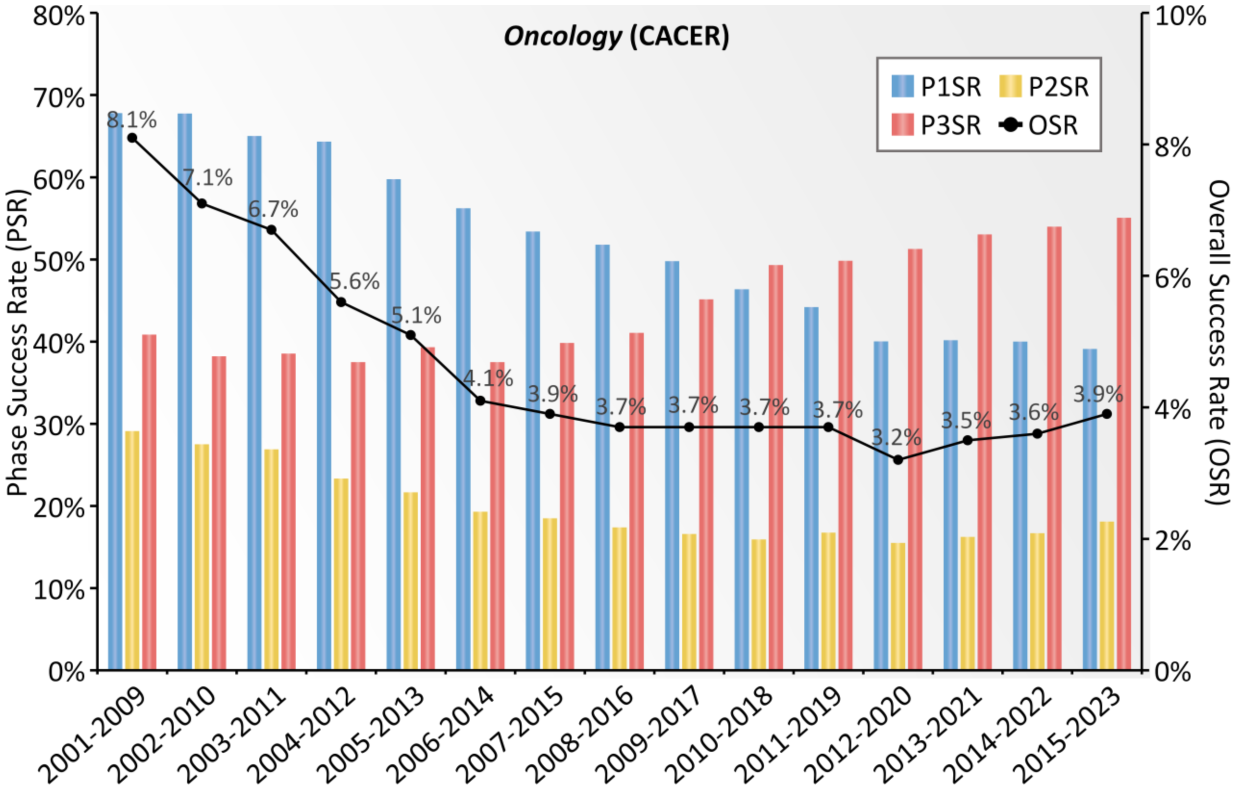


**Supplementary Figure S1**. The dynamic *clinical success rate* (ClinSR) evaluated based on the CTPs of *oncology* (CACER) collected for this study. A nine-year time-window was adopted here to facilitate the assessments of ClinSRs, which provided a drug adequate period of time to reach its final fate, and a total of fifteen time-windows (from 2001-2009 to 2015-2023, inclusive) were then assessed. The *phase success rates* (PSRs) of P1SR, P2SR and P3SR were shown using bars in blue, yellow and red, respectively. The dark line gave the dynamic variation in *overall success rate* (OSR). P1SR: Phase 1 success rate; P2SR: Phase 2 success rate; P3SR: Phase 3 success rate.

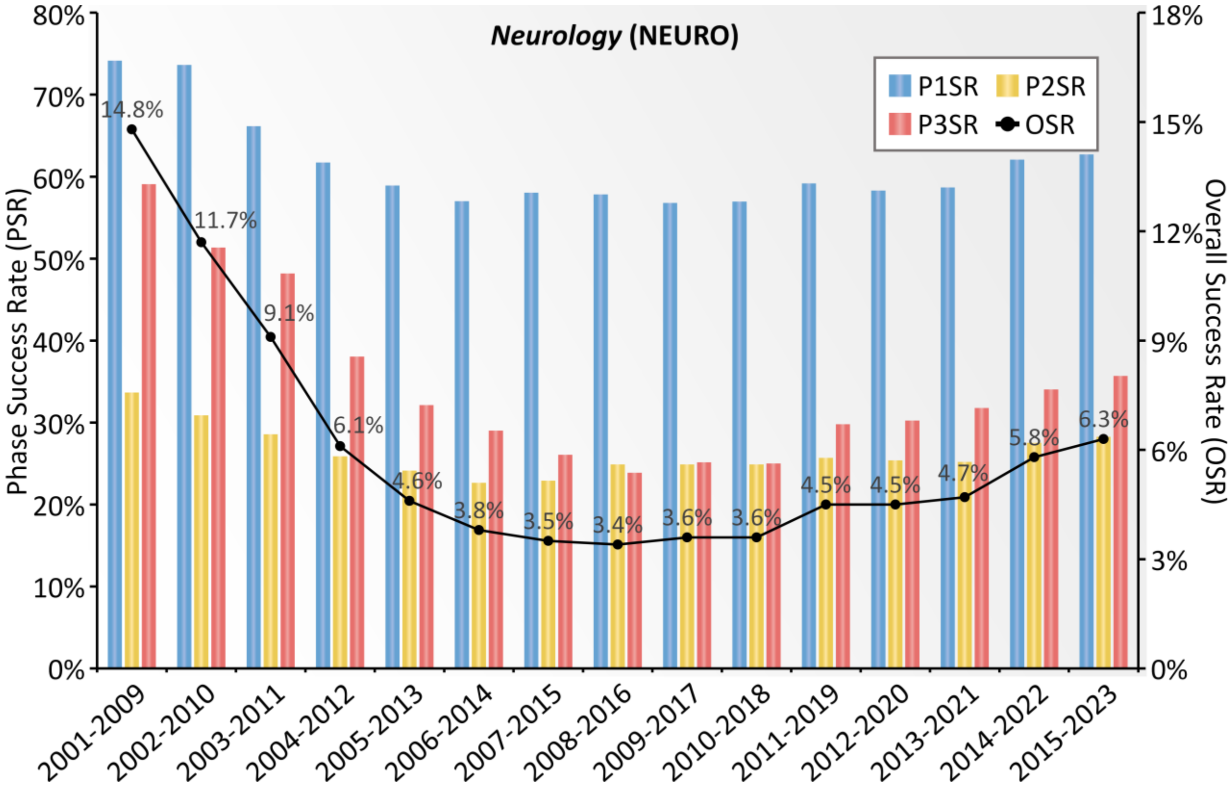


**Supplementary Figure S2**. Dynamic *clinical success rate* (ClinSR) assessed using the CTPs of *neurological disease* (NEURO) collected by this study. A nine-year time-window was used here to facilitate the assessments of ClinSRs, which provided a drug adequate period of time to reach its final fate, and a total of fifteen time-windows (from 2001-2009 to 2015-2023, inclusive) were then assessed. The *phase success rates* (PSRs) of P1SR, P2SR and P3SR were shown using bars in blue, yellow and red, respectively. The dark line gave the dynamic variation in *overall success rate* (OSR). P1SR: Phase 1 success rate; P2SR: Phase 2 success rate; P3SR: Phase 3 success rate.

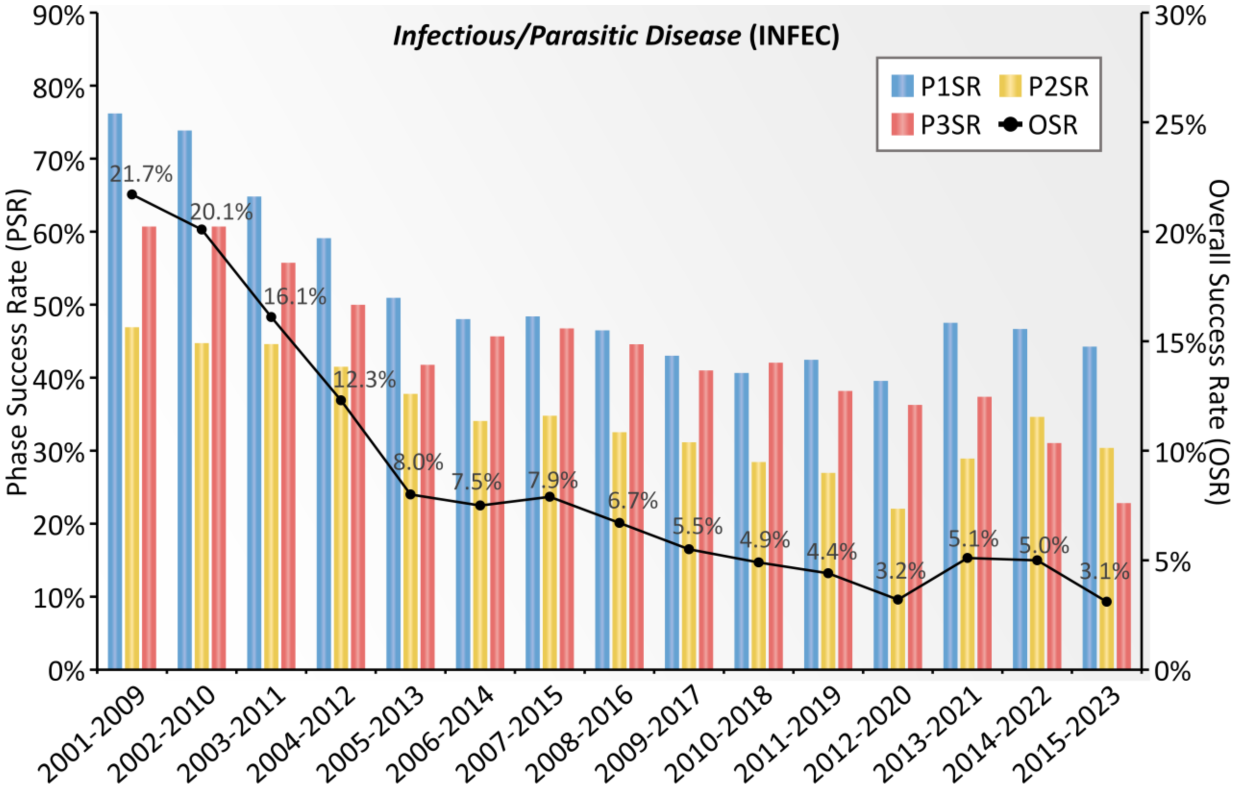


**Supplementary Figure S3**. Dynamic *clinical success rate* (ClinSR) assessed using the CTPs of *infectious/parasitic disease* (INFEC) collected for this study. A nine-year time-window was used to facilitate the assessments of ClinSRs, which provided a drug adequate period of time to reach its final fate, and a total of fifteen time-windows (from 2001-2009 to 2015-2023, inclusive) were then assessed. The *phase success rates* (PSRs) of P1SR, P2SR and P3SR were shown using bars in blue, yellow and red, respectively. The dark line gave the dynamic variation in *overall success rate* (OSR). P1SR: Phase 1 success rate; P2SR: Phase 2 success rate; P3SR: Phase 3 success rate.

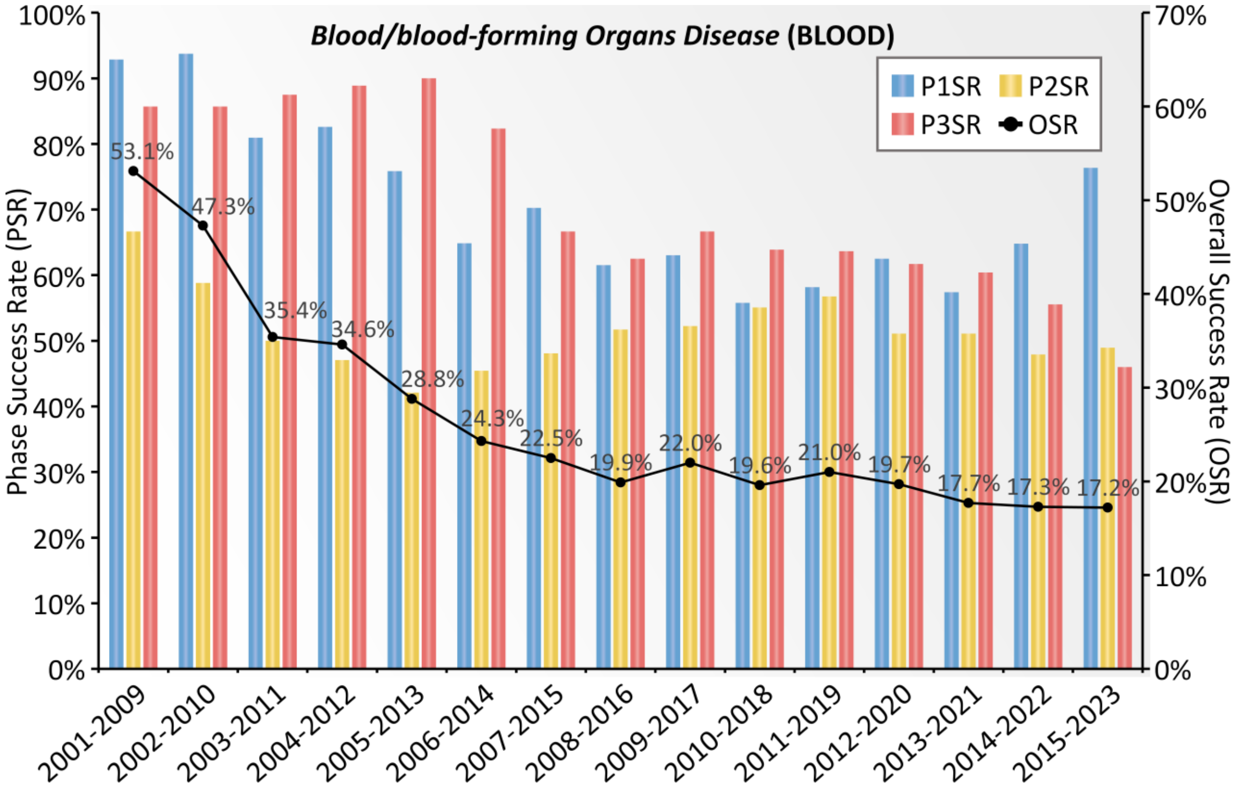


**Supplementary Figure S4**. Dynamic *clinical success rate* (ClinSR) assessed using the CTPs of *blood/blood-forming organs disease* (BLOOD) in this study. A nine-year time-window was used to facilitate the assessments of ClinSRs, which provided a drug adequate period of time to reach its final fate, and a total of fifteen time-windows (from 2001-2009 to 2015-2023, inclusive) were then assessed. The *phase success rates* (PSRs) of P1SR, P2SR and P3SR were shown using bars in blue, yellow and red, respectively. The dark line gave the dynamic variation in *overall success rate* (OSR). P1SR: Phase 1 success rate; P2SR: Phase 2 success rate; P3SR: Phase 3 success rate.

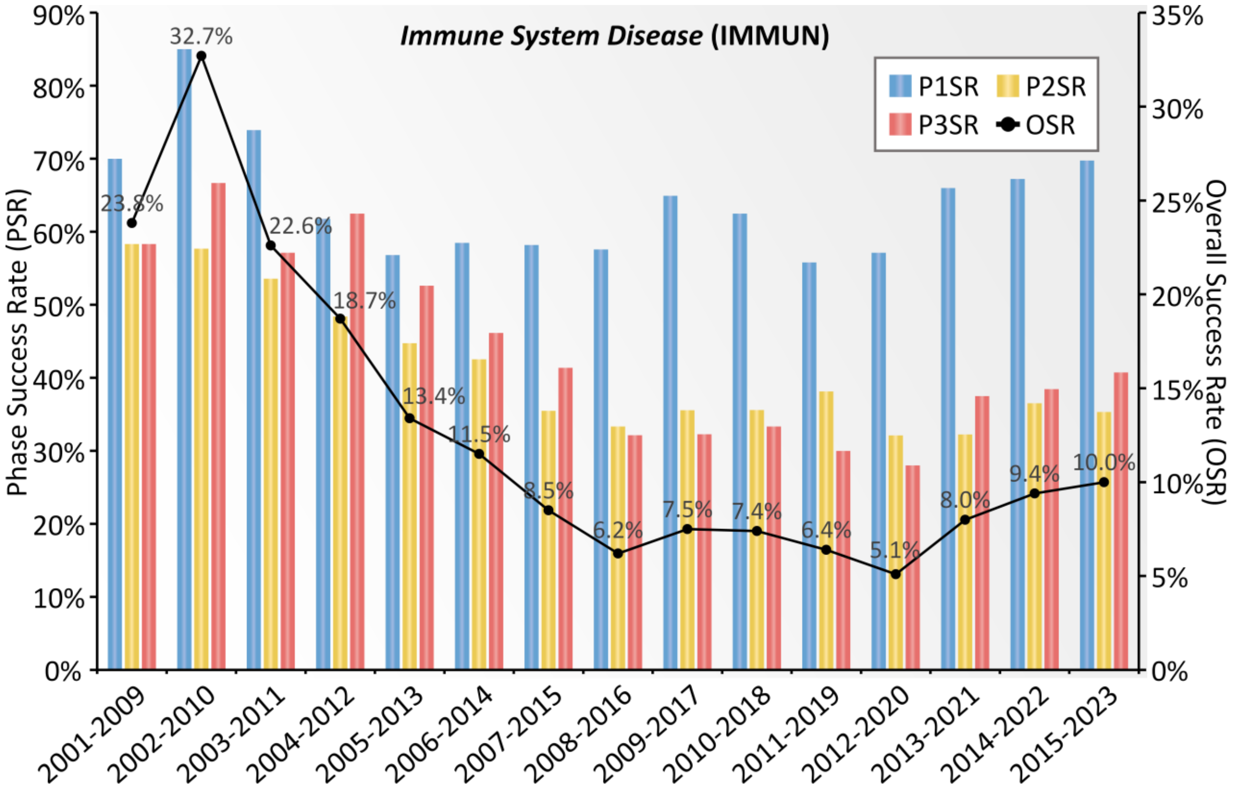


**Supplementary Figure S5**. Dynamic *clinical success rate* (ClinSR) assessed using the CTPs of *immune system disease* (IMMNU) collected by this study. A nine-year time-window was adopted to facilitate the assessments of ClinSRs, which provided a drug adequate period of time to reach its final fate, and a total of fifteen time-windows (from 2001-2009 to 2015-2023, inclusive) were then assessed. The *phase success rates* (PSRs) of P1SR, P2SR and P3SR were shown using bars in blue, yellow and red, respectively. The dark line gave the dynamic variation in *overall success rate* (OSR). P1SR: Phase 1 success rate; P2SR: Phase 2 success rate; P3SR: Phase 3 success rate.

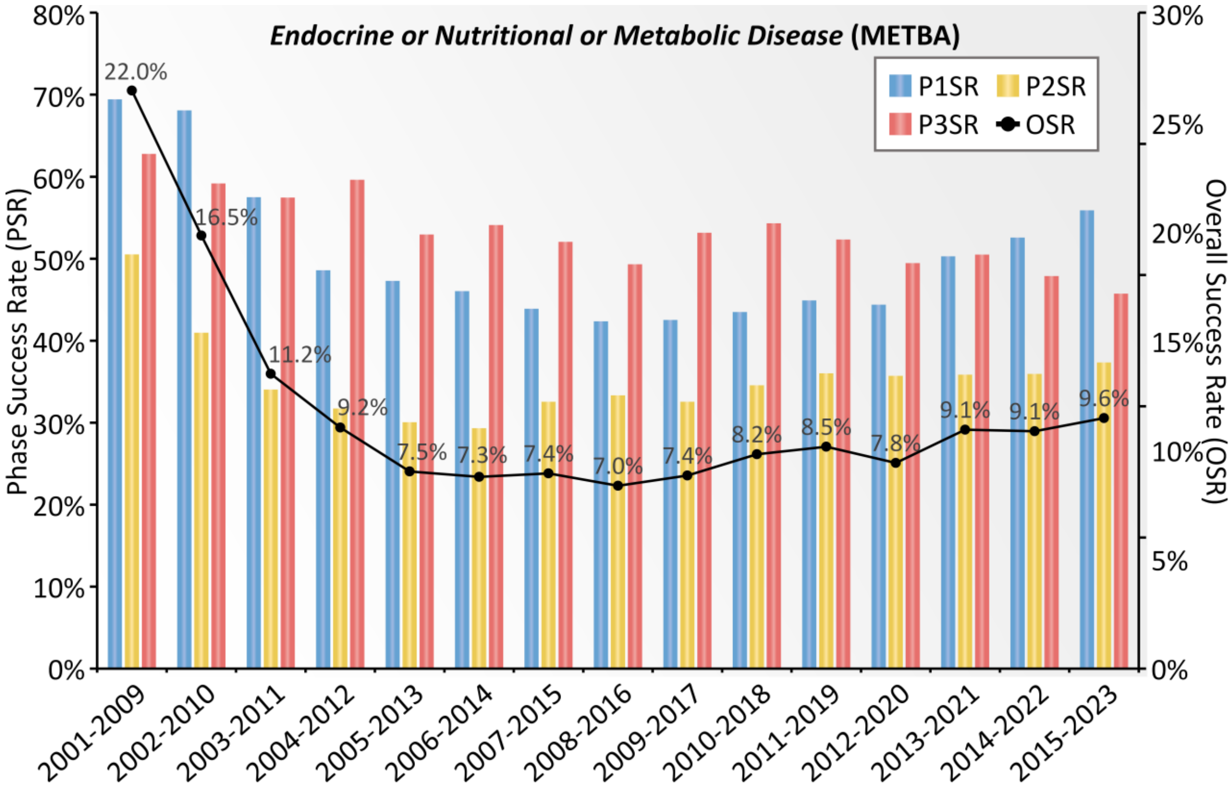


**Supplementary Figure S6**. Dynamic *clinical success rate* (ClinSR) assessed using the CTPs of *endocrine, nutritional or metabolic disease* (METAB). A nine-year time-window was used here to facilitate the assessments of ClinSRs, which provided a drug adequate period of time to reach its final fate, and a total of fifteen time-windows (from 2001-2009 to 2015-2023, inclusive) were then assessed. The *phase success rates* (PSRs) of P1SR, P2SR and P3SR were shown using bars in blue, yellow and red, respectively. The dark line gave the dynamic variation in *overall success rate* (OSR). P1SR: Phase 1 success rate; P2SR: Phase 2 success rate; P3SR: Phase 3 success rate.

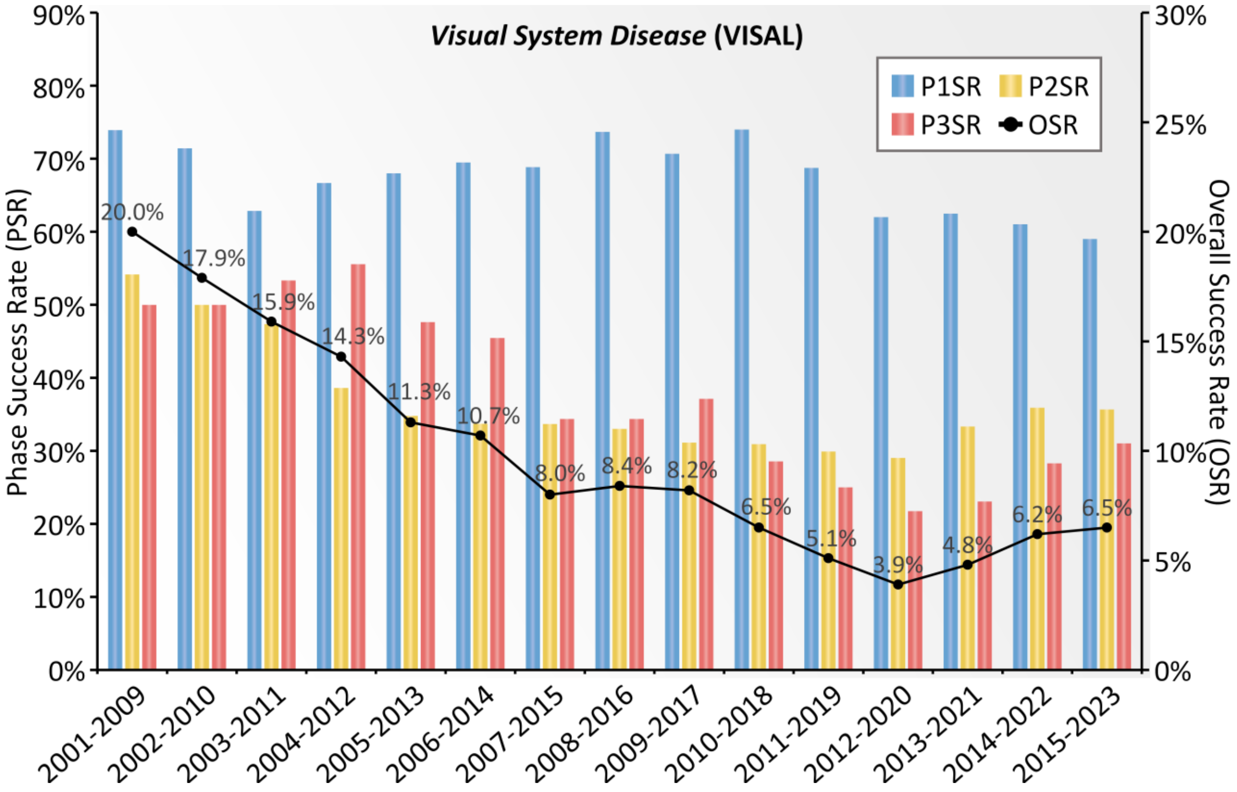


**Supplementary Figure S7**. Dynamic *clinical success rate* (ClinSR) assessed using the CTPs of *visual system disease* (VISAL) collected for this study. A nine-year time-window was used here to facilitate the assessments of ClinSRs, which provided a drug adequate period of time to reach its final fate, and a total of fifteen time-windows (from 2001-2009 to 2015-2023, inclusive) were then assessed. The *phase success rates* (PSRs) of P1SR, P2SR and P3SR were shown using bars in blue, yellow and red, respectively. The dark line gave the dynamic variation in *overall success rate* (OSR). P1SR: Phase 1 success rate; P2SR: Phase 2 success rate; P3SR: Phase 3 success rate.

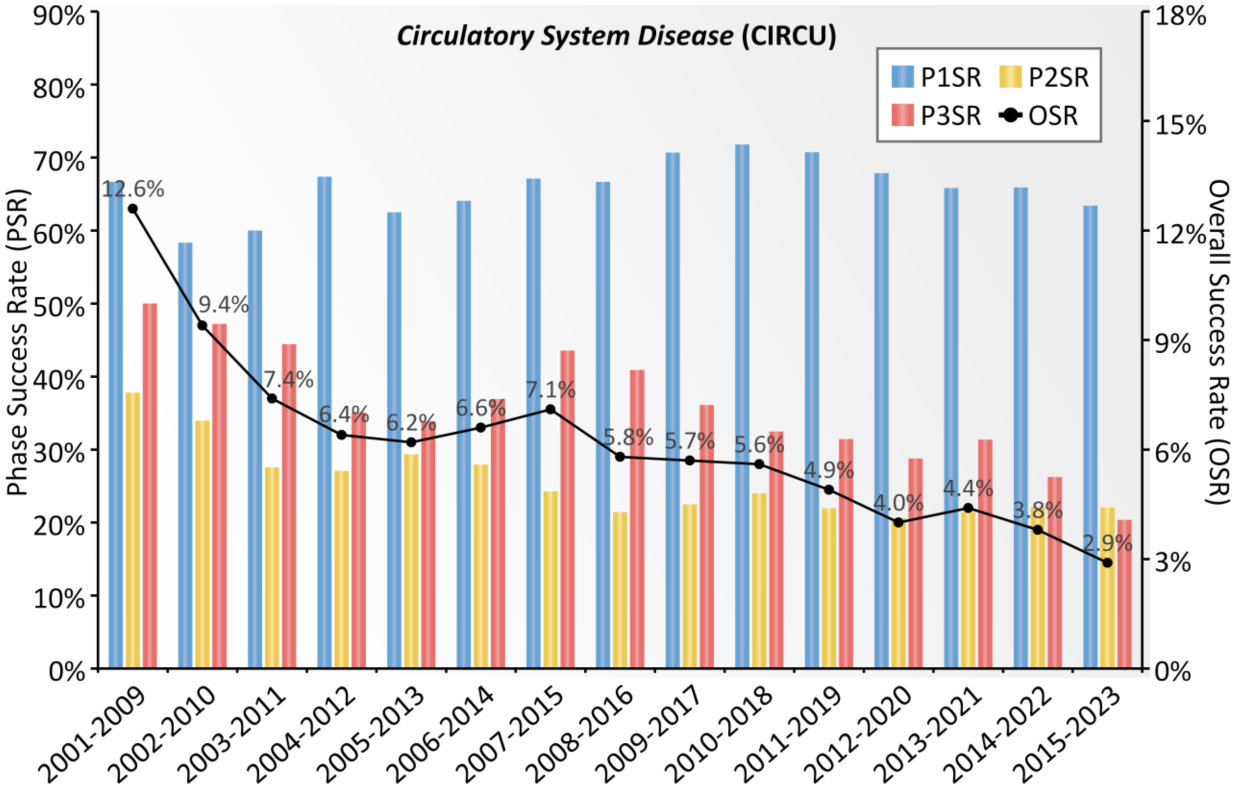


**Supplementary Figure S8**. Dynamic *clinical success rate* (ClinSR) assessed using the CTPs of *circulatory system disease* (CIRCU) collected for this study. A nine-year time-window was used to facilitate the assessments of ClinSRs, which provided a drug adequate period of time to reach its final fate, and a total of fifteen time-windows (from 2001-2009 to 2015-2023, inclusive) were then assessed. The *phase success rates* (PSRs) of P1SR, P2SR and P3SR were shown using bars in blue, yellow and red, respectively. The dark line gave the dynamic variation in *overall success rate* (OSR). P1SR: Phase 1 success rate; P2SR: Phase 2 success rate; P3SR: Phase 3 success rate.

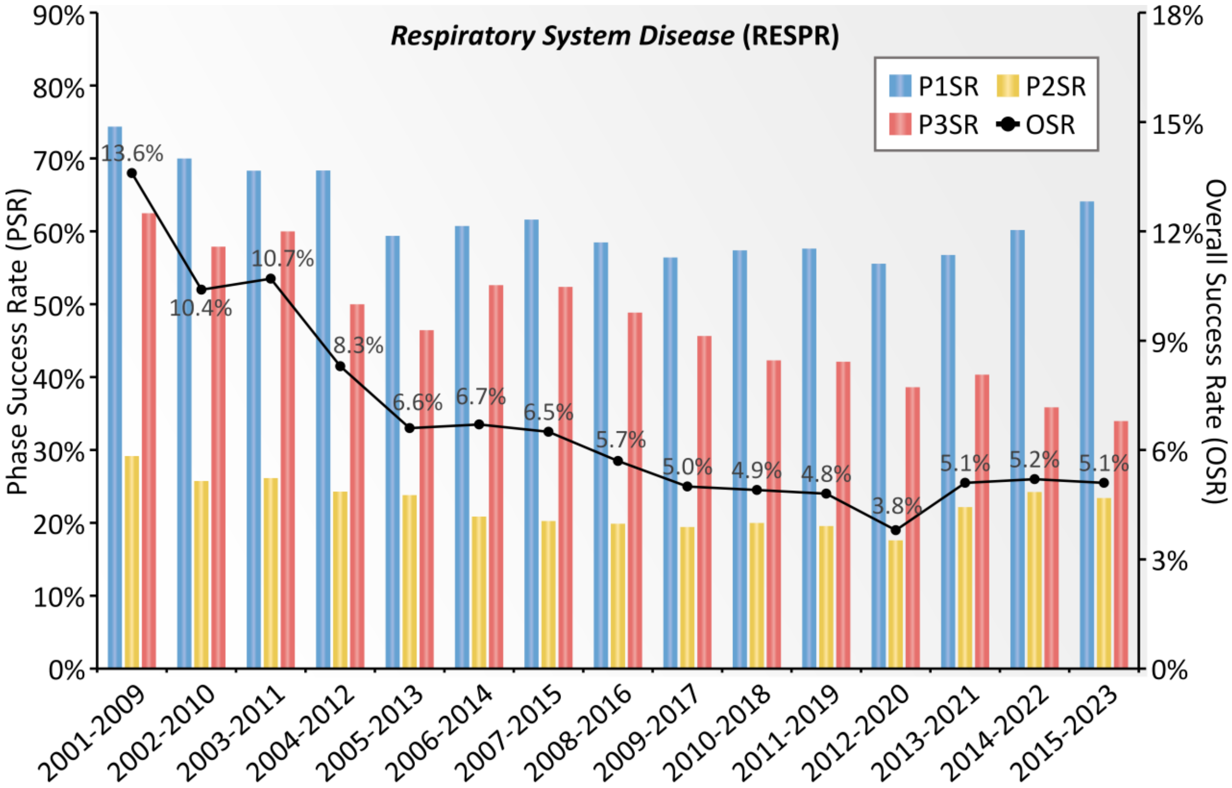


**Supplementary Figure S9**. Dynamic *clinical success rate* (ClinSR) assessed using the CTPs of *respiratory system disease* (RESPR) collected for this study. A nine-year time-window was used to facilitate the assessments of ClinSRs, which provided a drug adequate period of time to reach its final fate, and a total of fifteen time-windows (from 2001-2009 to 2015-2023, inclusive) were then assessed. The *phase success rates* (PSRs) of P1SR, P2SR and P3SR were shown using bars in blue, yellow and red, respectively. The dark line gave the dynamic variation in *overall success rate* (OSR). P1SR: Phase 1 success rate; P2SR: Phase 2 success rate; P3SR: Phase 3 success rate.

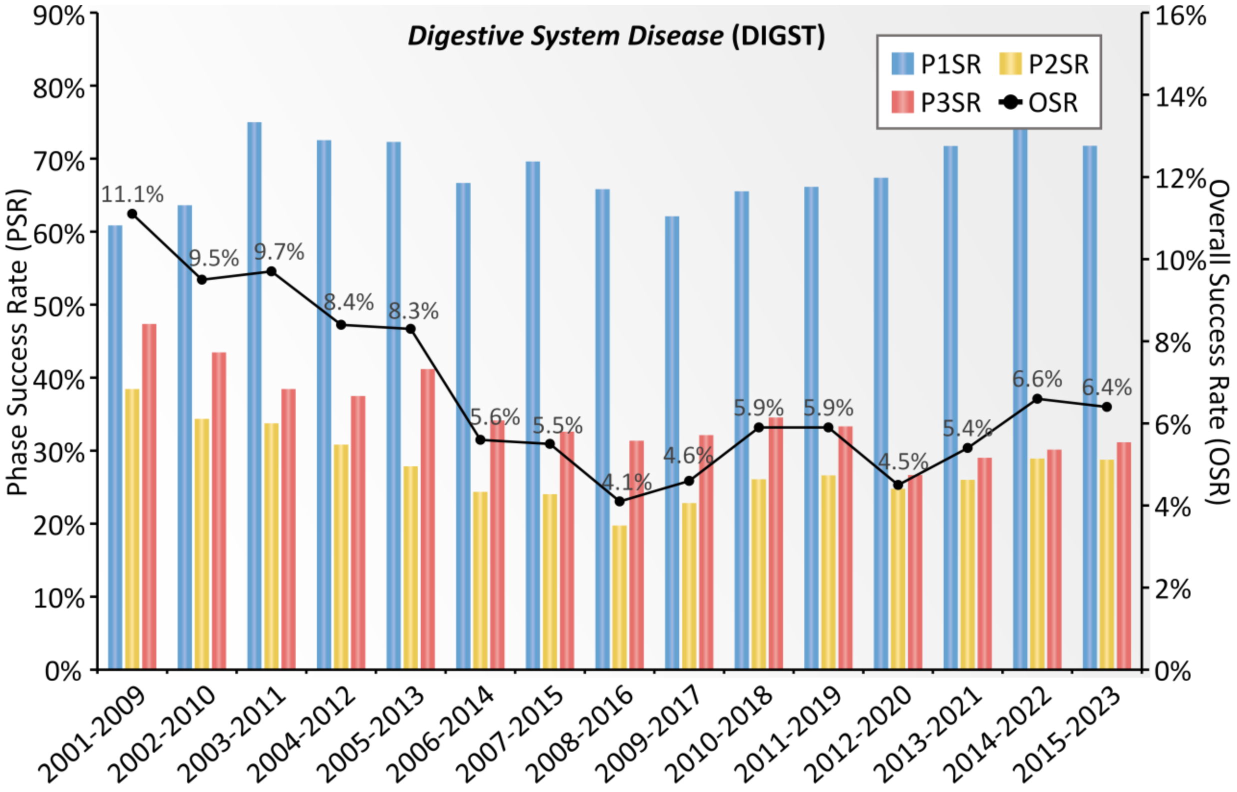


**Supplementary Figure S10**. Dynamic *clinical success rate* (ClinSR) evaluated using the CTPs of *digestive system disease* (DIGST) collected for this study. A nine-year time-window was used to facilitate the assessments of ClinSRs, which provided a drug adequate period of time to reach its final fate, and a total of fifteen time-windows (from 2001-2009 to 2015-2023, inclusive) were then assessed. The *phase success rates* (PSRs) of P1SR, P2SR and P3SR were shown using bars in blue, yellow and red, respectively. The dark line gave the dynamic variation in *overall success rate* (OSR). P1SR: Phase 1 success rate; P2SR: Phase 2 success rate; P3SR: Phase 3 success rate.

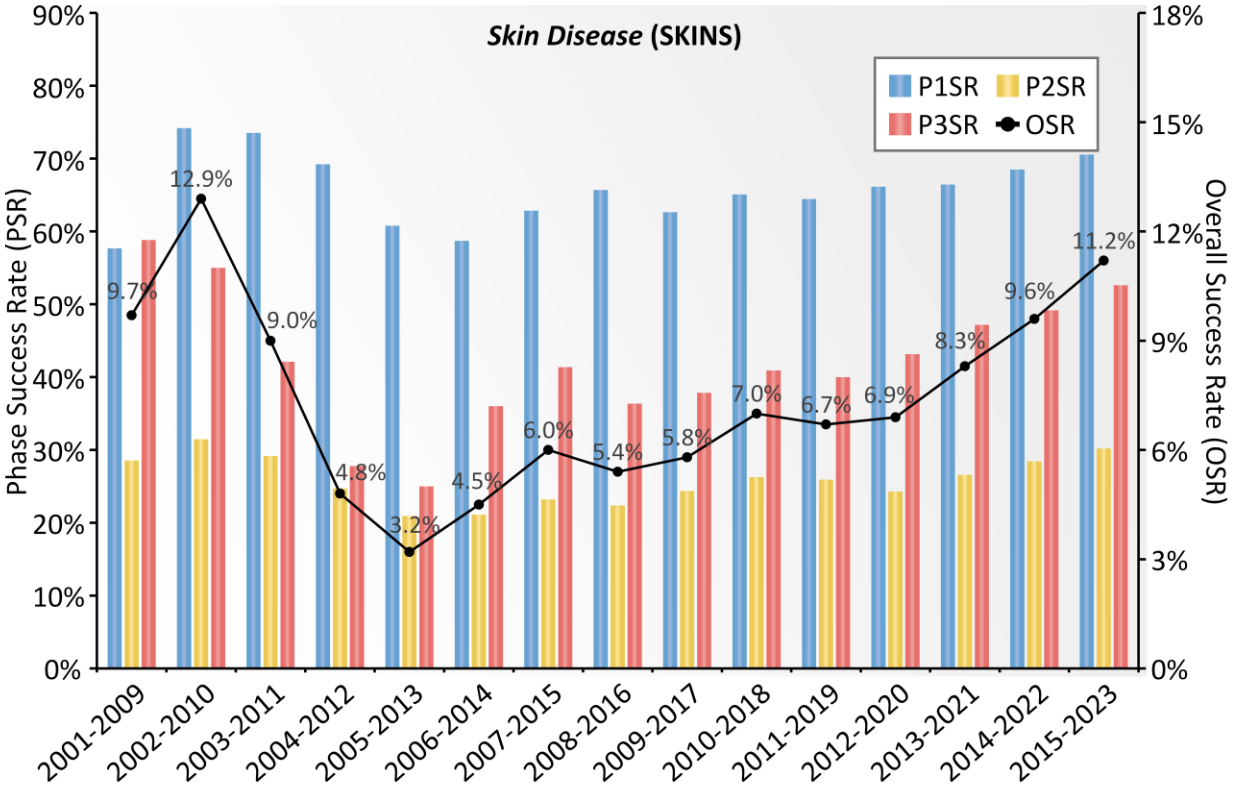


**Supplementary Figure S11**. The dynamic *clinical success rate* (ClinSR) assessed based on the CTPs of *skin disease* (SKINS) gathered in this study. A nine-year time-window was adopted here to facilitate the assessments of ClinSRs, which provided a drug adequate period of time to reach its final fate, and a total of fifteen time-windows (from 2001-2009 to 2015-2023, inclusive) were then assessed. The *phase success rates* (PSRs) of P1SR, P2SR and P3SR were shown using bars in blue, yellow and red, respectively. The dark line gave the dynamic variation in *overall success rate* (OSR). P1SR: Phase 1 success rate; P2SR: Phase 2 success rate; P3SR: Phase 3 success rate.

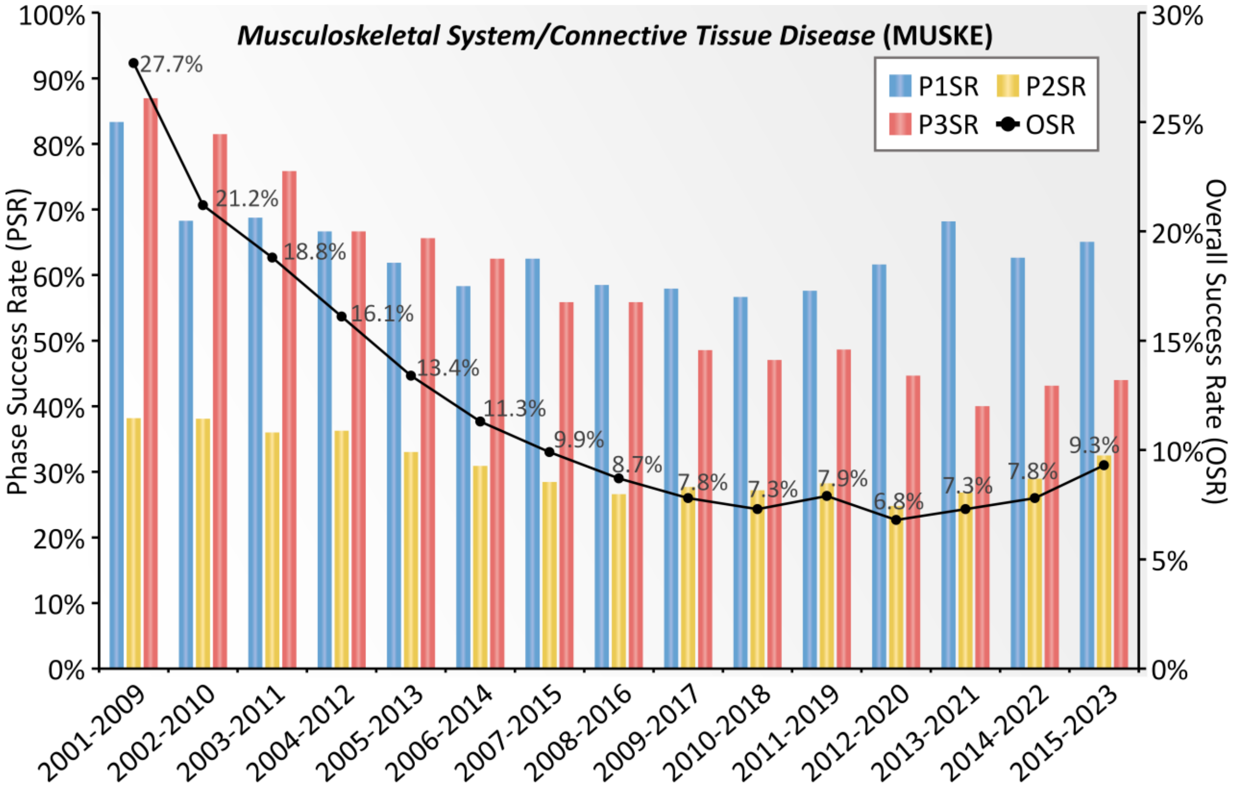


**Supplementary Figure S12**. Dynamic *clinical success rate* (ClinSR) measured by the CTPs of *musculoskeletal system/connective tissue disease* (MUSKE). A nine-year time-window was used to facilitate the assessments of ClinSRs, which provided a drug adequate period of time to reach its final fate, and a total of fifteen time-windows (from 2001-2009 to 2015-2023, inclusive) were then assessed. The *phase success rates* (PSRs) of P1SR, P2SR and P3SR were shown using bars in blue, yellow and red, respectively. The dark line gave the dynamic variation in *overall success rate* (OSR). P1SR: Phase 1 success rate; P2SR: Phase 2 success rate; P3SR: Phase 3 success rate.

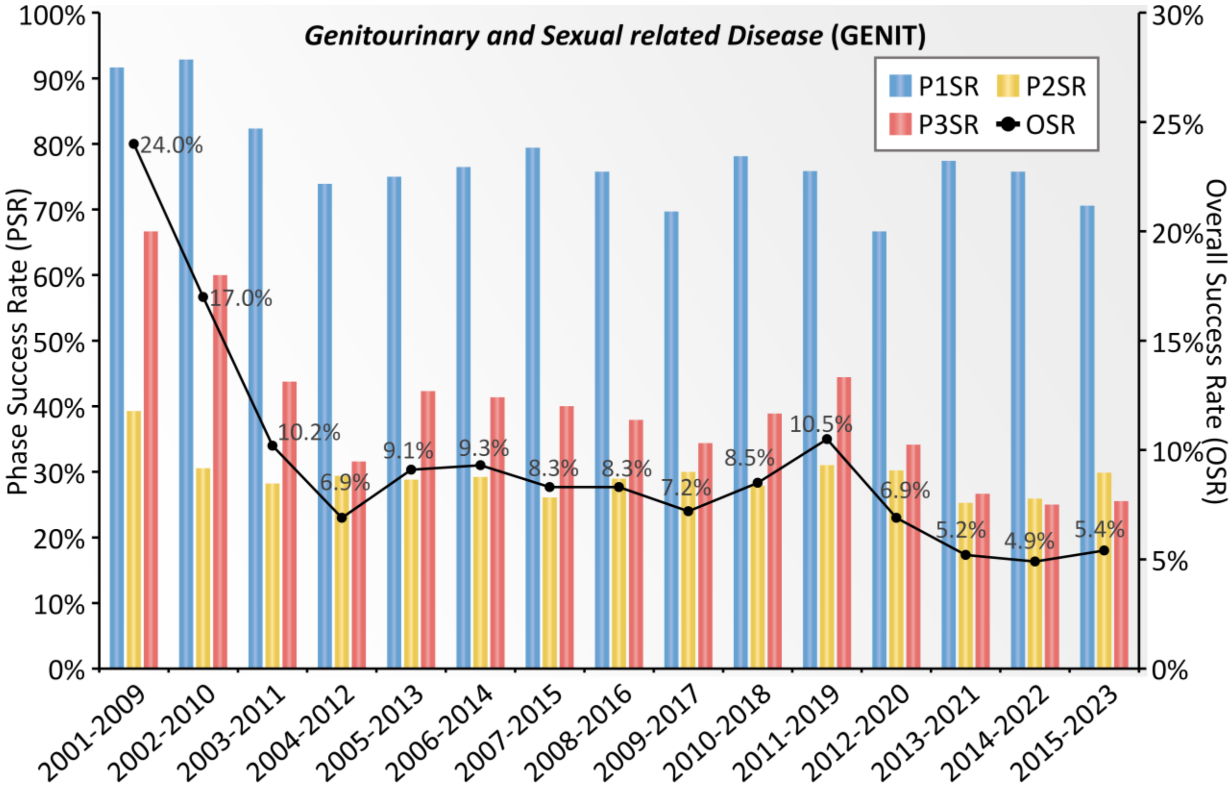


**Supplementary Figure S13**. Dynamic *clinical success rate* (ClinSR) measured by the CTPs of *genitourinary and sexual related disease* (GENIT). A nine-year time-window was adopted here to facilitate the assessments of ClinSRs, which provided a drug adequate period of time to reach its final fate, and a total of fifteen time-windows (from 2001-2009 to 2015-2023, inclusive) were then assessed. The *phase success rates* (PSRs) of P1SR, P2SR and P3SR were shown using bars in blue, yellow and red, respectively. The dark line gave the dynamic variation in *overall success rate* (OSR). P1SR: Phase 1 success rate; P2SR: Phase 2 success rate; P3SR: Phase 3 success rate.

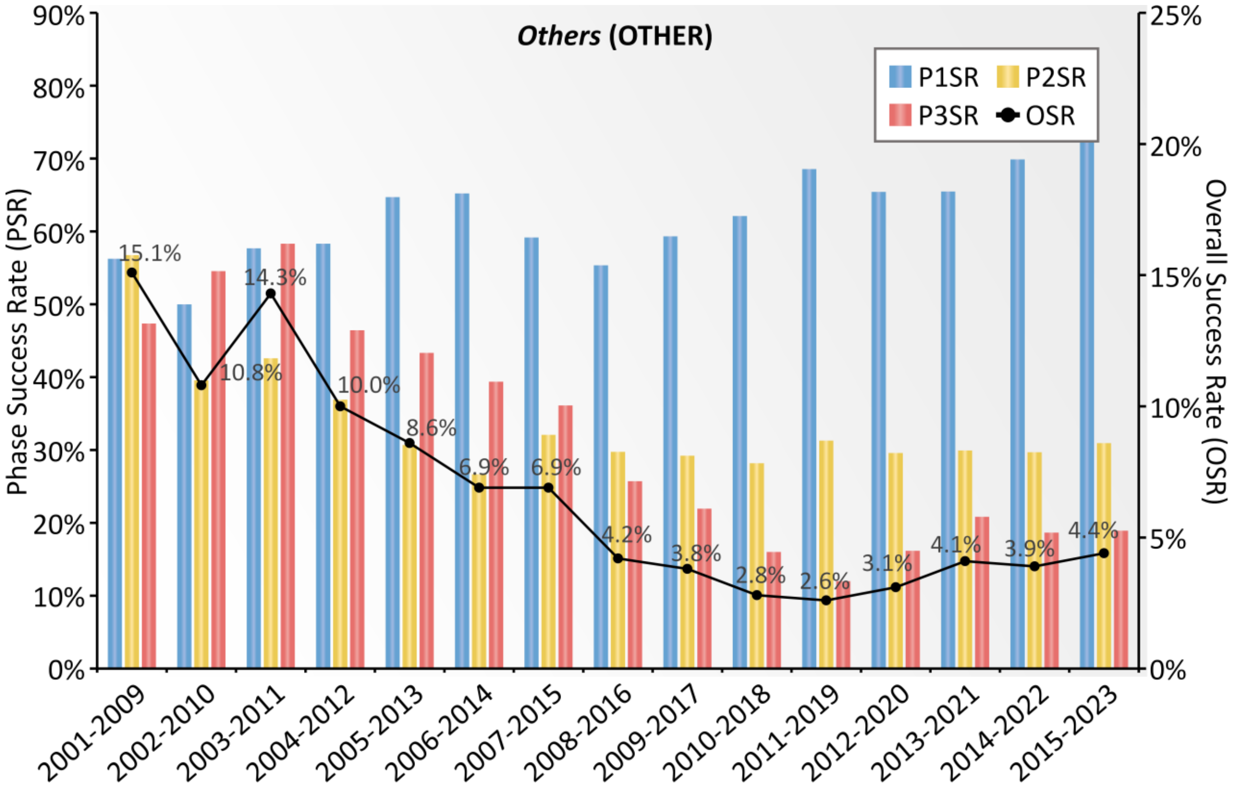


**Supplementary Figure S14**. The dynamic *clinical success rate* (ClinSR) measured by the CTPs of *other diseases* (OTHER) collected for this study. A nine-year time-window was adopted here to facilitate the assessments of ClinSRs, which provided a drug adequate period of time to reach its final fate, and a total of fifteen time-windows (from 2001-2009 to 2015-2023, inclusive) were then assessed. The *phase success rates* (PSRs) of P1SR, P2SR and P3SR were shown using bars in blue, yellow and red, respectively. The dark line gave the dynamic variation in *overall success rate* (OSR). P1SR: Phase 1 success rate; P2SR: Phase 2 success rate; P3SR: Phase 3 success rate.

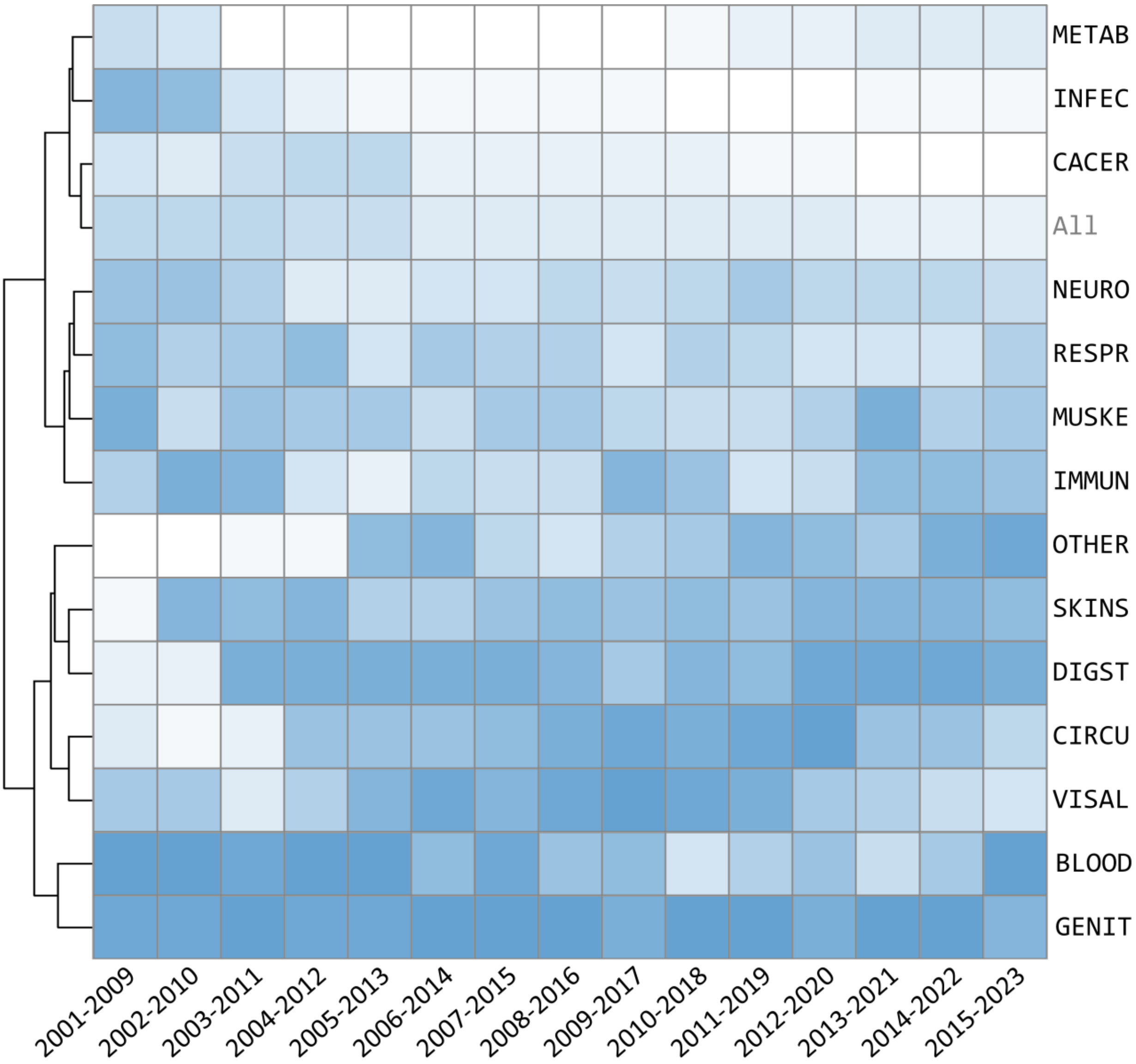


**Supplementary Figure S15**. Hierarchical clustering of fifteen different disease classes based on their *phase 1 success rate* (P1SR) across fifteen consecutive time-windows. The darker the blue color, the higher the success rate of clinical drug development in the corresponding time-window and the corresponding indication. All: all disease indication; INFEC: infectious/parasitic disease; BLOOD: blood/blood-forming organs disease; CACER: oncology; CIRCU: circulatory system disease; DIGST: digestive system disease; SKINS: skin disease; METAB: endocrine, nutritional or metabolic disease; RESPR: respiratory system disease; MUSKE: musculoskeletal system and connective tissue disease; GENIT: genitourinary and sexual related disease; NEURO: neurology; VISAL: visual system disease; IMMUN: immune system disease; OTHER: Other disease.

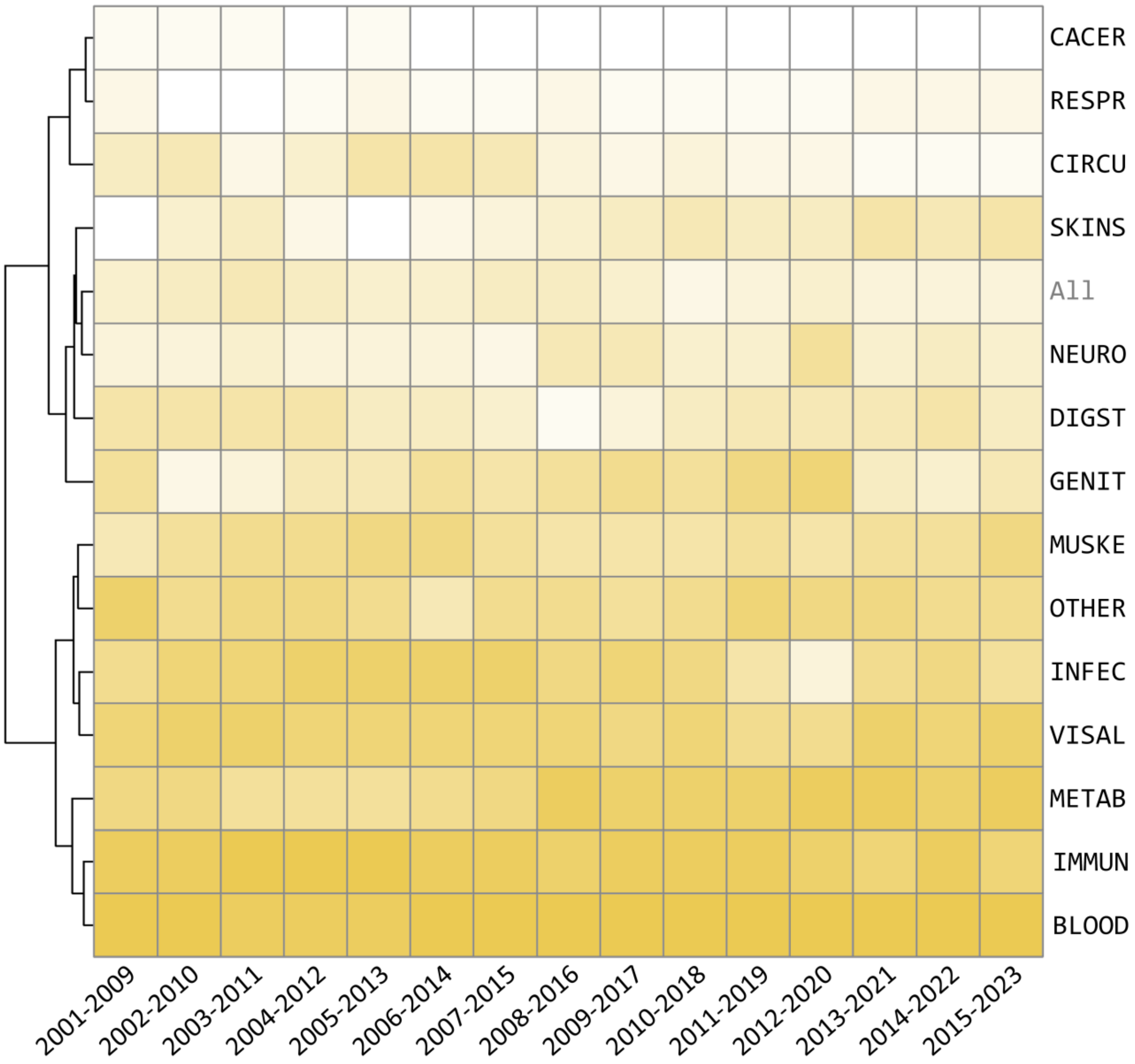


**Supplementary Figure S16**. Hierarchical clustering of fifteen different disease classes based on the *phase* 2 *success rate* (P2SR) across fifteen consecutive time-windows. The darker the yellow color, the higher the success rate of clinical drug development in the corresponding time-window and the corresponding indication. All: all disease indication; INFEC: infectious/parasitic disease; BLOOD: blood/blood-forming organs disease; CACER: oncology; CIRCU: circulatory system disease; DIGST: digestive system disease; SKINS: skin disease; METAB: endocrine, nutritional or metabolic disease; RESPR: respiratory system disease; MUSKE: musculoskeletal system and connective tissue disease; GENIT: genitourinary and sexual related disease; NEURO: neurology; VISAL: visual system disease; IMMUN: immune system disease; OTHER: Other disease.

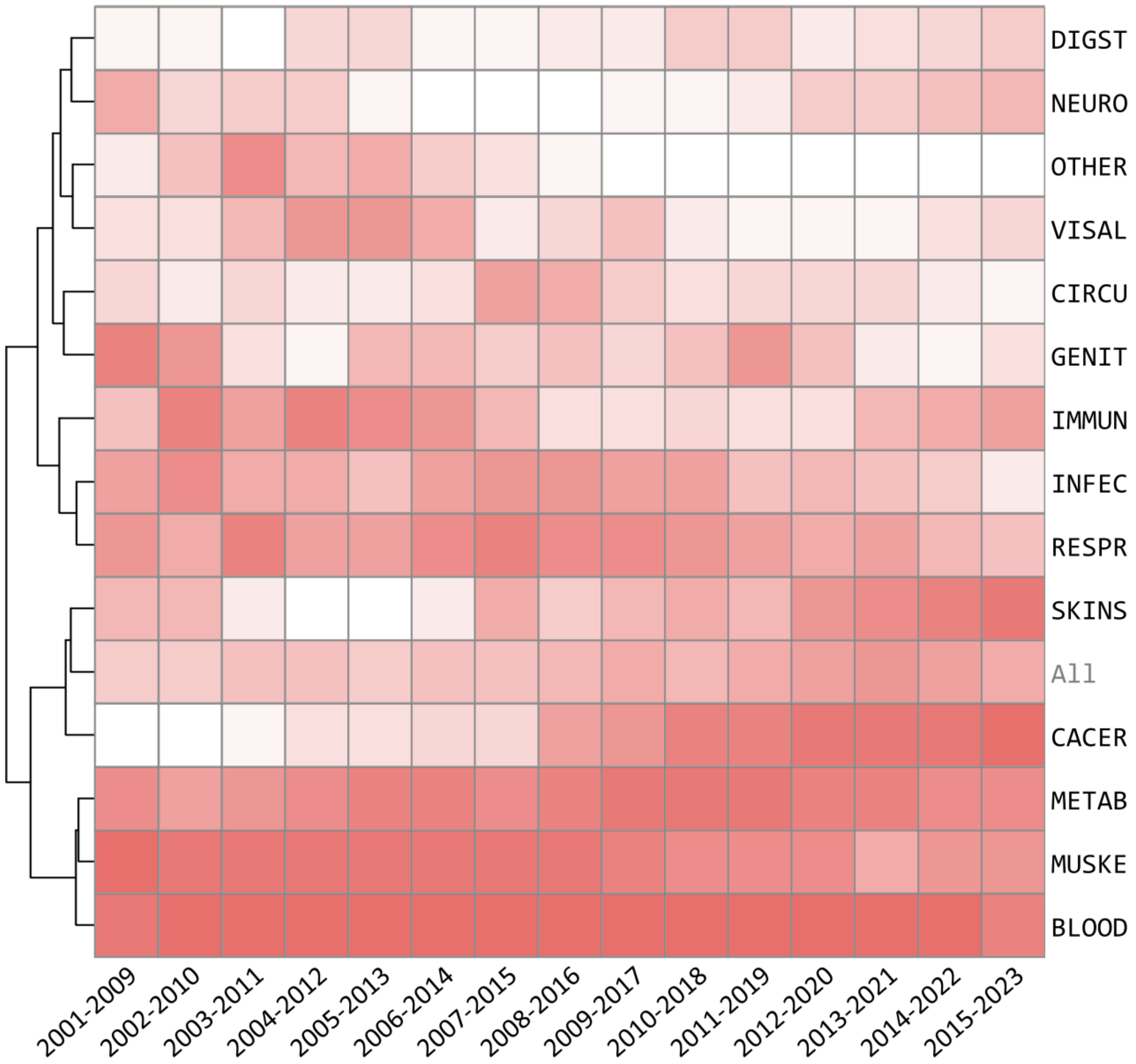


**Supplementary Figure S17**. Hierarchical clustering of fifteen different disease classes based on their *phase 3 success rate* (P3SR) across fifteen consecutive time-windows. The darker the red color, the higher the success rate of clinical drug development in the corresponding time-window and the corresponding indication. All: all disease indication; INFEC: infectious/parasitic disease; BLOOD: blood/blood-forming organs disease; CACER: oncology; CIRCU: circulatory system disease; DIGST: digestive system disease; SKINS: skin disease; METAB: endocrine, nutritional or metabolic disease; RESPR: respiratory system disease; MUSKE: musculoskeletal system and connective tissue disease; GENIT: genitourinary and sexual related disease; NEURO: neurology; VISAL: visual system disease; IMMUN: immune system disease; OTHER: Other disease.

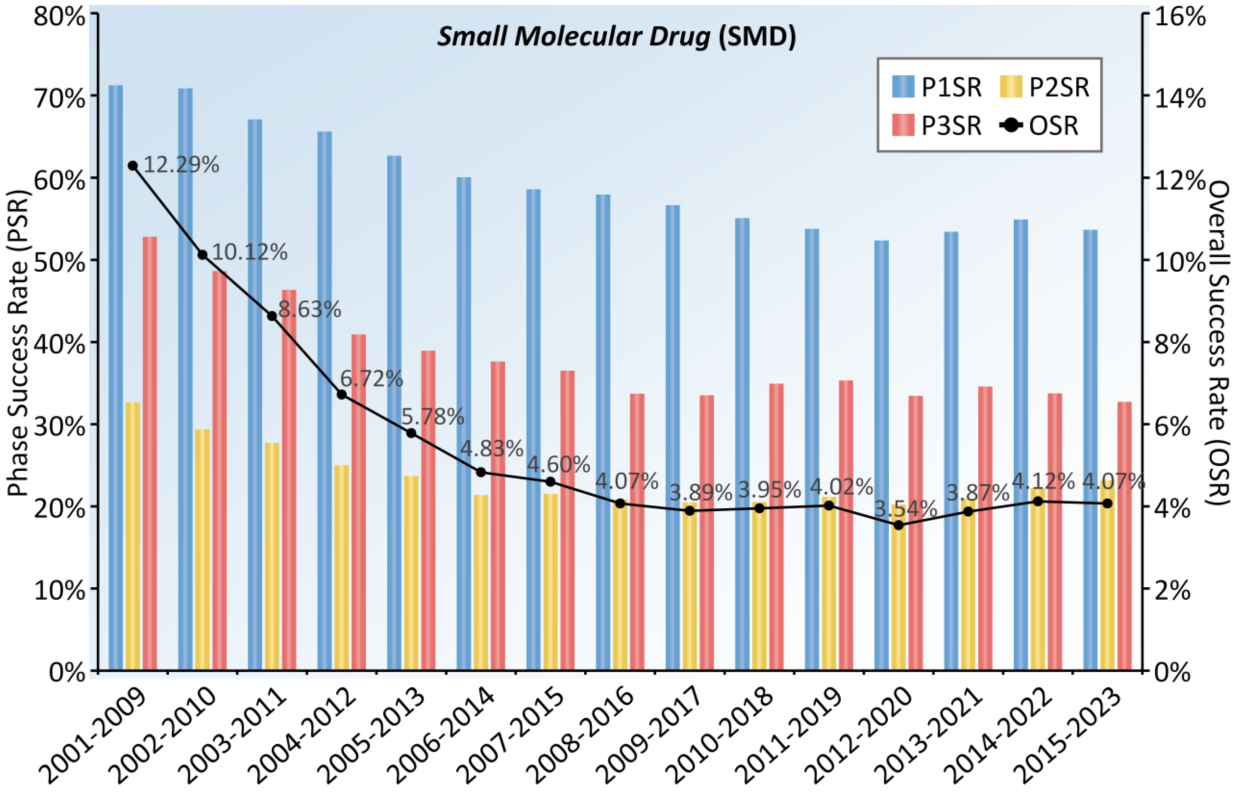


**Supplementary Figure S18**. The dynamic *clinical success rate* (ClinSR) measured by the CTPs of *small molecular drug* (SMD) collected for this study. A nine-year time-window was used here to facilitate the assessments of ClinSRs, which provided a drug adequate period of time to reach its final fate, and a total of fifteen time-windows (from 2001-2009 to 2015-2023, inclusive) were then assessed. The *phase success rates* (PSRs) of P1SR, P2SR and P3SR were shown using bars in blue, yellow and red, respectively. The dark line gave the dynamic variation in *overall success rate* (OSR). P1SR: Phase 1 success rate; P2SR: Phase 2 success rate; P3SR: Phase 3 success rate.

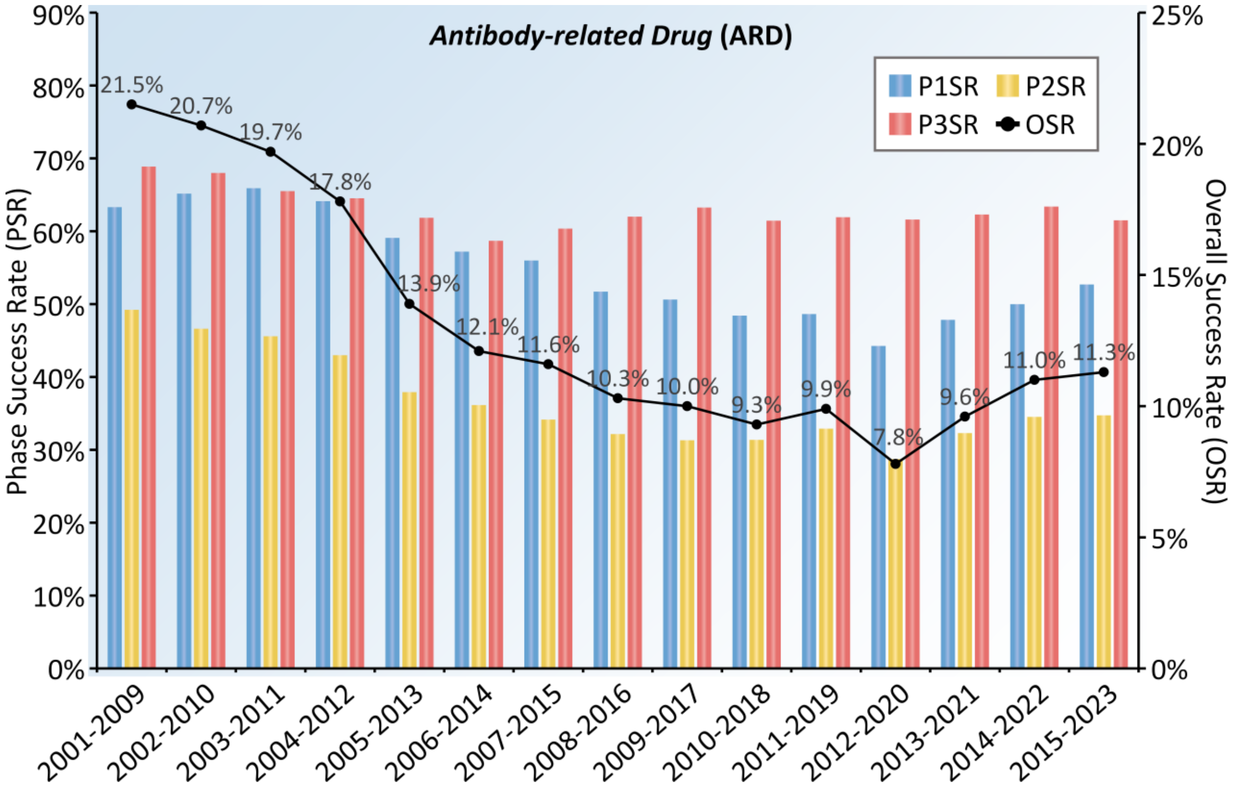


**Supplementary Figure S19**. The dynamic *clinical success rate* (ClinSR) measured by the CTPs of *antibody-related drug* (ARD) collected for this study. A nine-year time-window was used here to facilitate the assessments of ClinSRs, which provided a drug adequate period of time to reach its final fate, and a total of fifteen time-windows (from 2001-2009 to 2015-2023, inclusive) were then assessed. The *phase success rates* (PSRs) of P1SR, P2SR and P3SR were shown using bars in blue, yellow and red, respectively. The dark line gave the dynamic variation in *overall success rate* (OSR). P1SR: Phase 1 success rate; P2SR: Phase 2 success rate; P3SR: Phase 3 success rate.

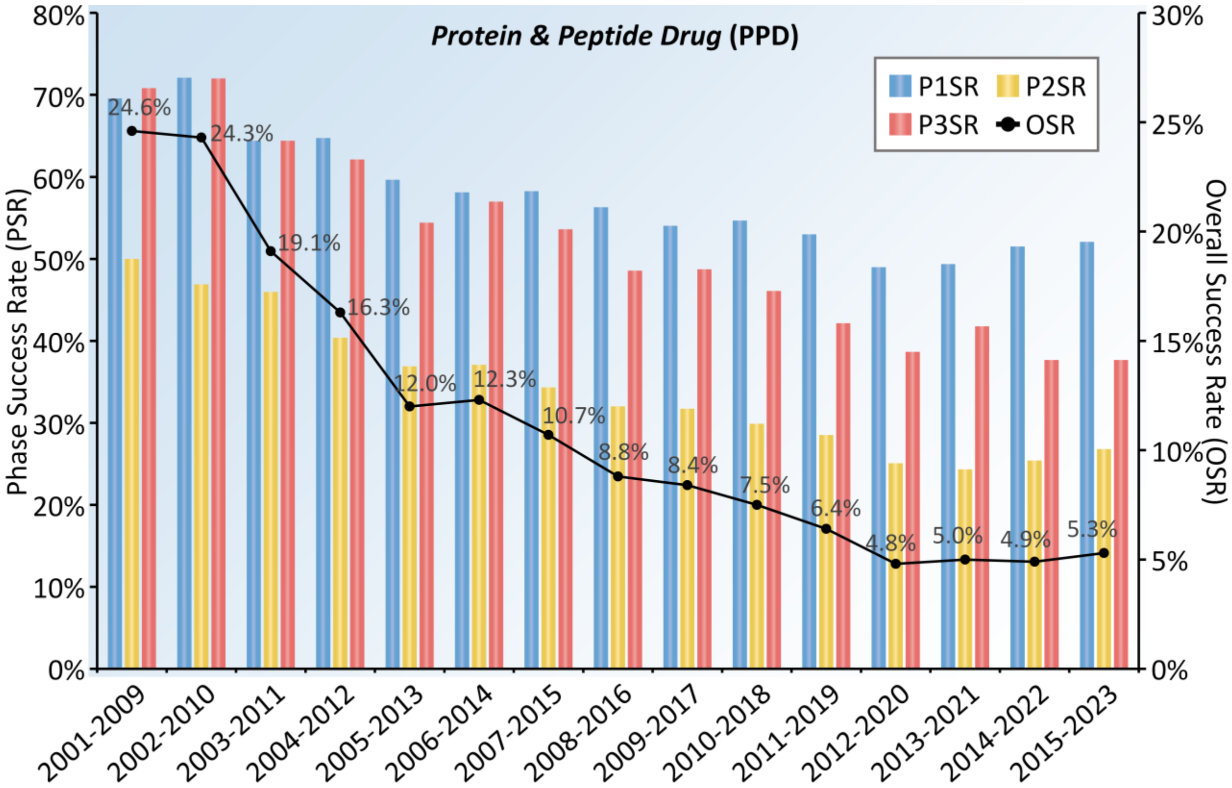


**Supplementary Figure S20**. The dynamic *clinical success rate* (ClinSR) measured by the CTPs of *protein & peptide drug* (PPD) collected for this study. A nine-year time-window was used here to facilitate the assessments of ClinSRs, which provided a drug adequate period of time to reach its final fate, and a total of fifteen time-windows (from 2001-2009 to 2015-2023, inclusive) were then assessed. The *phase success rates* (PSRs) of P1SR, P2SR and P3SR were shown using bars in blue, yellow and red, respectively. The dark line gave the dynamic variation in *overall success rate* (OSR). P1SR: Phase 1 success rate; P2SR: Phase 2 success rate; P3SR: Phase 3 success rate.

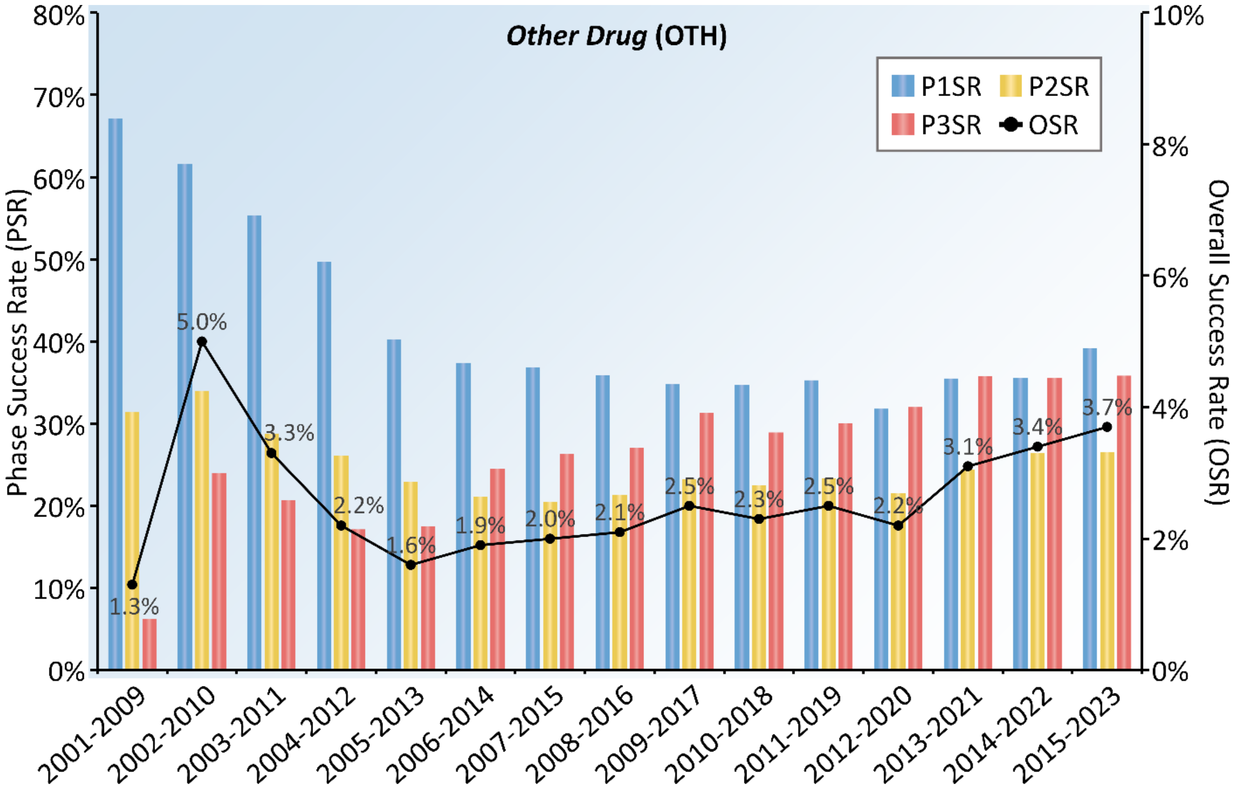


**Supplementary Figure S21**. The dynamic *clinical success rate* (ClinSR) assessed based on the CTPs of *other drug* (OTH) collected for this study. A nine-year time-window was adopted here to facilitate the assessments of ClinSRs, which provided a drug adequate period of time to reach its final fate, and a total of fifteen time-windows (from 2001-2009 to 2015-2023, inclusive) were then assessed. The *phase success rates* (PSRs) of P1SR, P2SR and P3SR were shown using bars in blue, yellow and red, respectively. The dark line gave the dynamic variation in *overall success rate* (OSR). P1SR: Phase 1 success rate; P2SR: Phase 2 success rate; P3SR: Phase 3 success rate.

**Supplementary Table S1**. The representative studies previously published that related to the analysis of *clinical success rate* (ClinSR) of drugs.

| | |
|---|---|
| **(*a*) Representative studies analyzing the success rates for the whole industry** | |
| Approval success rates of drug candidates based on target, action, modality, and their combinations | *Clin Transl Sci*. 2021; 14: 1113 |
| Trends in clinical success rates and therapeutic focus | *Nat Rev Drug Discov*. 2019; 18: 495 |
| Estimation of clinical trial success rates and related parameters | *Biostatistics*. 2019; 20: 273 |
| Trends in clinical success rates | *Nat Rev Drug Discov*. 2016; 15: 379 |
| Clinical development success rates for investigational drugs | *Nat Biotechnol*. 2014; 32: 40 |
| Trends in risks associated with new drug development success rates for investigational drugs | *Clin Pharmacol Ther*. 2010; 87: 272 |
| Can the pharmaceutical industry reduce attrition rates | *Nat Rev Drug Discov*. 2004; 3: 711 |
| **(*b*) Representative studies analyzing the success rates for certain disease class** | |
| Development times and approval success rates for drugs to treat **infectious diseases** | *Clin Pharmacol Ther*. 2020; 107: 324 |
| Changes in clinical trials of **cancer** drugs in mainland China over the decade 2009-18 | *Lancet Oncol*. 2019; 20: e619 |
| Lessons learned from two decades of **anticancer** drugs | *Trends Pharmacol Sci*. 2017; 38: 852 |
| A quantitative analysis of therapeutic **cancer vaccines** in phase 2 or phase 3 trial | *J Immunother Cancer*. 2015; 3:48 |
| Clinical approval success rates for investigational **cancer** drugs | *Clin Pharmacol Ther*. 2013; 94: 329 |
| Development trends for new **cancer** therapeutics and vaccines | *Drug Discov Today*. 2008; 13: 30 |
| **(*c*) Representative studies analyzing the success rates for certain individual disease** | |
| The wider perspective twenty years of clinical trials in **myelodysplastic syndromes** | *Br J Haematol*. 2022; 196: 329 |

| | |
|---|---|
| **Alzheimer's** disease key insights from two decades of clinical trial failures | *J Alzheimers Dis*. 2022; 87 :83 |
| **Parkinson's** disease drug development since 1999 a story of repurposing and relative success | *J Parkinsons Dis*. 2021;11: 421 |
| Clinical trial risk in **leukemia** Biomarkers and trial design | *Hematol Oncol*. 2021; 39: 105 |
| Transition probabilities for clinical trials investigating **individual diseases** | *Nat Rev Drug Discov*. 2019; 18: 658 |
| Drug development for **breast, colorectal, and non-small cell lung cancers** from 1979 to 2014 | *Cancer*. 2017; 123: 4672 |
| Clinical trial risk in **chronic obstructive pulmonary disease** the effects of drug class and criteria | *Respiration*. 2016; 91: 79 |
| Reducing clinical trial risk in **multiple sclerosis** | *Mult Scler Relat Disord*. 2016; 5: 8 |
| Clinical trial risk in **hepatitis C** endpoint selection and drug action | *CJG Hepatol*. 2016; 2016: 6260271 |
| Clinical trial success rates of anti-**obesity** agents the importance of combination therapies | *Obes Rev*. 2015; 16: 707 |
| **Alzheimer's** disease drug-development pipeline few candidates, frequent failures | *Alzheimers Res Ther*. 2014; 6 :37 |
| Clinical trial risk in **Non-Hodgkin's lymphoma** endpoint and target selection | *J Pharm Pharm Sci*. 2011; 14: 227 |
| The success rate of new drug development in clinical trials **Crohn's disease** | *J Pharm Pharm Sci*. 2010;13: 191 |
| **(*d*) Representative studies analyzing the success rates for a certain company** | |
| Achieving end-to-end success in the clinic **Pfizer**'s learnings on R&D productivity | *Drug Discov Today*. 2022; 27: 697 |
| Reviving an R&D pipeline a step change in the Phase II success rate | *Drug Discov Today*. 2021; 26: 308 |

**Supplementary Table S2**. The *phase 1 success rate*s (P1SRs) for all CTPs and the CTPs of particular disease class (a total of 14 classes defined by WHO ICD-11) calculated across fifteen nine-year time-windows. All: all diseases; INFEC: Infectious/parasitic disease; CACER: Oncology; BLOOD: Blood/blood-forming organs disease; IMMUN: Immune system disease; METAB: Endocrine, nutritional or metabolic disease; NEURO: Neurology; VISAL: Visual system disease; CIRCU: Circulatory system disease; RESPR: Respiratory system disease; DIGST: Digestive system disease; SKINS: Skin disease; MUSKE: Musculoskeletal system/connective tissue disease; GENIT: Genitourinary and sexual related disease; OTHER: Other disease.

| Disease Class | 2001 -2009 | 2002 -2010 | 2003 -2011 | 2004 -2012 | 2005 -2013 | 2006 -2014 | 2007 -2015 | 2008 -2016 | 2009 -2017 | 2010 -2018 | 2011 -2019 | 2012 -2020 | 2013 -2021 | 2014 -2022 | 2015 -2023 |
|---|---|---|---|---|---|---|---|---|---|---|---|---|---|---|---|
| All | 69.7% | 69.4% | 65.6% | 63.7% | 59.4% | 56.9% | 55.8% | 54.2% | 52.9% | 51.5% | 50.7% | 48.0% | 49.8% | 51.0% | 51.3% |
| 01 INFEC | 76.2% | 73.9% | 64.8% | 59.1% | 51.0% | 48.0% | 48.4% | 46.5% | 43.0% | 40.7% | 42.5% | 39.6% | 47.5% | 46.7% | 44.3% |
| 02 CACER | 67.8% | 67.8% | 65.0% | 64.3% | 59.8% | 56.2% | 53.4% | 51.8% | 49.8% | 46.4% | 44.2% | 40.0% | 40.2% | 40.0% | 39.1% |
| 03 BLOOD | 92.9% | 93.8% | 81.0% | 82.6% | 75.9% | 64.9% | 70.3% | 61.5% | 63.0% | 55.8% | 58.2% | 62.5% | 57.4% | 64.8% | 76.4% |
| 04 IMMUN | 70.0% | 85.0% | 73.9% | 61.8% | 56.8% | 58.5% | 58.2% | 57.6% | 64.9% | 62.5% | 55.8% | 57.1% | 66.0% | 67.2% | 69.8% |
| 05 METAB | 69.4% | 68.1% | 57.5% | 48.6% | 47.3% | 46.1% | 43.9% | 42.4% | 42.5% | 43.5% | 44.9% | 44.4% | 50.3% | 52.6% | 55.9% |
| 06 NEURO | 74.2% | 73.6% | 66.2% | 61.7% | 58.9% | 57.0% | 58.1% | 57.8% | 56.8% | 57.0% | 59.2% | 58.3% | 58.7% | 62.1% | 62.7% |
| 07 VISAL | 73.9% | 71.4% | 62.9% | 66.7% | 68.0% | 69.5% | 68.9% | 73.7% | 70.7% | 74.0% | 68.8% | 62.0% | 62.5% | 61.0% | 59.0% |
| 08 CIRCU | 66.7% | 58.3% | 60.0% | 67.4% | 62.5% | 64.1% | 67.1% | 66.7% | 70.7% | 71.8% | 70.7% | 67.9% | 65.8% | 65.9% | 63.4% |
| 09 RESPR | 74.4% | 70.0% | 68.3% | 68.4% | 59.4% | 60.8% | 61.6% | 58.5% | 56.4% | 57.4% | 57.6% | 55.6% | 56.8% | 60.2% | 64.1% |

| | | | | | | | | | | | | | | | |
|---|---|---|---|---|---|---|---|---|---|---|---|---|---|---|---|
| 10 DIGST | 60.9% | 63.6% | 75.0% | 72.6% | 72.3% | 66.7% | 69.6% | 65.8% | 62.1% | 65.6% | 66.1% | 67.4% | 71.7% | 75.2% | 71.8% |
| 11 SKINS | 57.7% | 74.2% | 73.5% | 69.2% | 60.8% | 58.7% | 62.9% | 65.7% | 62.6% | 65.1% | 64.5% | 66.1% | 66.4% | 68.5% | 70.6% |
| 12 MUSKE | 83.3% | 68.3% | 68.8% | 66.7% | 61.9% | 58.3% | 62.5% | 58.5% | 58.0% | 56.7% | 57.6% | 61.6% | 68.2% | 62.6% | 65.1% |
| 13 GENIT | 91.7% | 92.9% | 82.4% | 73.9% | 75.0% | 76.5% | 79.4% | 75.8% | 69.7% | 78.1% | 75.9% | 66.7% | 77.4% | 75.8% | 70.6% |
| 14 OTHER | 56.3% | 50.0% | 57.7% | 58.3% | 64.7% | 65.2% | 59.2% | 55.4% | 59.3% | 62.1% | 68.6% | 65.4% | 65.5% | 69.9% | 75.0% |

**Supplementary Table S3**. The *phase 2 success rates* (P2SRs) for all CTPs and the CTPs of particular disease class (a total of 14 classes defined by WHO ICD-11) calculated across fifteen nine-year time-windows. All: all diseases; INFEC: Infectious/parasitic disease; CACER: Oncology; BLOOD: Blood/blood-forming organs disease; IMMUN: Immune system disease; METAB: Endocrine, nutritional or metabolic disease; NEURO: Neurology; VISAL: Visual system disease; CIRCU: Circulatory system disease; RESPR: Respiratory system disease; DIGST: Digestive system disease; SKINS: Skin disease; MUSKE: Musculoskeletal system/connective tissue disease; GENIT: Genitourinary and sexual related disease; OTHER: Other disease.

| Disease Class | 2001 -2009 | 2002 -2010 | 2003 -2011 | 2004 -2012 | 2005 -2013 | 2006 -2014 | 2007 -2015 | 2008 -2016 | 2009 -2017 | 2010 -2018 | 2011 -2019 | 2012 -2020 | 2013 -2021 | 2014 -2022 | 2015 -2023 |
|---|---|---|---|---|---|---|---|---|---|---|---|---|---|---|---|
| All | 35.8% | 32.8% | 31.1% | 28.3% | 26.3% | 24.2% | 24.1% | 23.3% | 23.2% | 23.1% | 23.8% | 22.2% | 23.6% | 25.1% | 26.0% |
| 01 INFEC | 46.9% | 44.7% | 44.6% | 41.5% | 37.8% | 34.1% | 34.8% | 32.5% | 31.2% | 28.4% | 27.0% | 22.1% | 28.9% | 34.6% | 30.4% |
| 02 CACER | 29.1% | 27.5% | 26.9% | 23.3% | 21.7% | 19.3% | 18.5% | 17.4% | 16.6% | 15.9% | 16.8% | 15.5% | 16.2% | 16.7% | 18.1% |
| 03 BLOOD | 66.7% | 58.8% | 50.0% | 47.1% | 42.1% | 45.5% | 48.1% | 51.7% | 52.2% | 55.1% | 56.8% | 51.1% | 51.1% | 47.9% | 48.9% |
| 04 IMMUN | 58.3% | 57.7% | 53.6% | 48.4% | 44.7% | 42.6% | 35.5% | 33.3% | 35.6% | 35.6% | 38.1% | 32.1% | 32.2% | 36.5% | 35.3% |
| 05 METAB | 50.5% | 41.0% | 34.0% | 31.7% | 30.1% | 29.3% | 32.6% | 33.3% | 32.6% | 34.6% | 36.0% | 35.7% | 35.9% | 36.0% | 37.3% |
| 06 NEURO | 33.7% | 30.9% | 28.6% | 25.9% | 24.1% | 22.7% | 22.9% | 24.9% | 24.9% | 24.9% | 25.7% | 25.4% | 25.2% | 27.5% | 28.3% |
| 07 VISAL | 54.2% | 50.0% | 47.4% | 38.6% | 34.8% | 33.7% | 33.7% | 33.0% | 31.1% | 30.9% | 29.9% | 29.0% | 33.3% | 35.9% | 35.7% |
| 08 CIRCU | 37.8% | 34.0% | 27.6% | 27.1% | 29.4% | 28.0% | 24.3% | 21.4% | 22.5% | 24.0% | 22.0% | 20.6% | 21.4% | 22.1% | 22.1% |
| 09 RESPR | 29.2% | 25.8% | 26.1% | 24.3% | 23.8% | 20.9% | 20.3% | 19.9% | 19.4% | 20.0% | 19.6% | 17.6% | 22.2% | 24.2% | 23.4% |

| | | | | | | | | | | | | | | | |
|---|---|---|---|---|---|---|---|---|---|---|---|---|---|---|---|
| 10 DIGST | 38.5% | 34.4% | 33.8% | 30.9% | 27.9% | 24.4% | 24.1% | 19.7% | 22.8% | 26.1% | 26.6% | 24.8% | 26.0% | 28.9% | 28.8% |
| 11 SKINS | 28.6% | 31.5% | 29.2% | 24.7% | 21.0% | 21.1% | 23.2% | 22.4% | 24.4% | 26.3% | 25.9% | 24.3% | 26.6% | 28.5% | 30.2% |
| 12 MUSKE | 38.2% | 38.1% | 36.0% | 36.3% | 33.0% | 30.9% | 28.5% | 26.6% | 27.7% | 27.2% | 28.2% | 24.8% | 26.8% | 29.0% | 32.5% |
| 13 GENIT | 39.3% | 30.6% | 28.2% | 29.4% | 28.8% | 29.2% | 26.1% | 29.0% | 30.0% | 27.9% | 31.0% | 30.2% | 25.3% | 25.9% | 29.9% |
| 14 OTHER | 56.8% | 39.6% | 42.6% | 36.9% | 30.7% | 26.7% | 32.1% | 29.8% | 29.2% | 28.2% | 31.3% | 29.6% | 29.9% | 29.7% | 30.9% |

**Supplementary Table S4**. The *phase 3 success rate*s (P3SRs) for all CTPs and the CTPs of particular disease class (a total of 14 classes defined by WHO ICD-11) calculated across fifteen nine-year time-windows. All: all diseases; INFEC: Infectious/parasitic disease; CACER: Oncology; BLOOD: Blood/blood-forming organs disease; IMMUN: Immune system disease; METAB: Endocrine, nutritional or metabolic disease; NEURO: Neurology; VISAL: Visual system disease; CIRCU: Circulatory system disease; RESPR: Respiratory system disease; DIGST: Digestive system disease; SKINS: Skin disease; MUSKE: Musculoskeletal system/connective tissue disease; GENIT: Genitourinary and sexual related disease; OTHER: Other disease.

| Disease Class | 2001 -2009 | 2002 -2010 | 2003 -2011 | 2004 -2012 | 2005 -2013 | 2006 -2014 | 2007 -2015 | 2008 -2016 | 2009 -2017 | 2010 -2018 | 2011 -2019 | 2012 -2020 | 2013 -2021 | 2014 -2022 | 2015 -2023 |
|---|---|---|---|---|---|---|---|---|---|---|---|---|---|---|---|
| All | 54.6% | 51.5% | 48.8% | 44.0% | 41.6% | 41.2% | 40.7% | 38.8% | 39.4% | 40.0% | 40.1% | 39.0% | 40.8% | 40.2% | 39.2% |
| 01 INFEC | 60.7% | 60.7% | 55.7% | 50.0% | 41.8% | 45.7% | 46.8% | 44.6% | 41.0% | 42.1% | 38.2% | 36.3% | 37.4% | 31.1% | 22.8% |
| 02 CACER | 40.9% | 38.2% | 38.6% | 37.5% | 39.3% | 37.5% | 39.8% | 41.1% | 45.2% | 49.3% | 49.8% | 51.3% | 53.0% | 54.0% | 55.1% |
| 03 BLOOD | 85.7% | 85.7% | 87.5% | 88.9% | 90.0% | 82.4% | 66.7% | 62.5% | 66.7% | 63.9% | 63.6% | 61.7% | 60.4% | 55.6% | 46.0% |
| 04 IMMUN | 58.3% | 66.7% | 57.1% | 62.5% | 52.6% | 46.2% | 41.4% | 32.1% | 32.3% | 33.3% | 30.0% | 28.0% | 37.5% | 38.5% | 40.7% |
| 05 METAB | 62.8% | 59.2% | 57.5% | 59.6% | 52.9% | 54.1% | 52.1% | 49.3% | 53.2% | 54.3% | 52.3% | 49.5% | 50.5% | 47.9% | 45.7% |
| 06 NEURO | 59.1% | 51.4% | 48.2% | 38.0% | 32.1% | 29.0% | 26.1% | 23.9% | 25.2% | 25.0% | 29.8% | 30.2% | 31.8% | 34.1% | 35.7% |
| 07 VISAL | 50.0% | 50.0% | 53.3% | 55.6% | 47.6% | 45.5% | 34.4% | 34.4% | 37.1% | 28.6% | 25.0% | 21.7% | 23.1% | 28.3% | 31.0% |
| 08 CIRCU | 50.0% | 47.2% | 44.4% | 35.1% | 33.9% | 36.9% | 43.6% | 40.9% | 36.1% | 32.5% | 31.4% | 28.8% | 31.3% | 26.2% | 20.4% |
| 09 RESPR | 62.5% | 57.9% | 60.0% | 50.0% | 46.4% | 52.6% | 52.4% | 48.8% | 45.7% | 42.3% | 42.1% | 38.6% | 40.4% | 35.9% | 34.0% |

| | | | | | | | | | | | | | | | |
|---|---|---|---|---|---|---|---|---|---|---|---|---|---|---|---|
| 10 DIGST | 47.4% | 43.5% | 38.5% | 37.5% | 41.2% | 34.2% | 32.6% | 31.4% | 32.1% | 34.6% | 33.3% | 26.7% | 29.0% | 30.2% | 31.2% |
| 11 SKINS | 58.8% | 55.0% | 42.1% | 27.8% | 25.0% | 36.0% | 41.4% | 36.4% | 37.8% | 40.9% | 40.0% | 43.1% | 47.2% | 49.2% | 52.6% |
| 12 MUSKE | 87.0% | 81.5% | 75.9% | 66.7% | 65.6% | 62.5% | 55.9% | 55.9% | 48.6% | 47.1% | 48.7% | 44.7% | 40.0% | 43.1% | 44.0% |
| 13 GENIT | 66.7% | 60.0% | 43.8% | 31.6% | 42.3% | 41.4% | 40.0% | 37.9% | 34.4% | 38.9% | 44.4% | 34.2% | 26.7% | 25.0% | 25.5% |
| 14 OTHER | 47.4% | 54.6% | 58.3% | 46.4% | 43.3% | 39.4% | 36.1% | 25.7% | 22.0% | 16.0% | 12.0% | 16.2% | 20.8% | 18.7% | 18.9% |
